# Patterns of antibiotic use, pathogens and clinical outcomes in hospitalised neonates and young infants with sepsis in the NeoOBS global neonatal sepsis observational cohort study

**DOI:** 10.1101/2022.06.20.22276674

**Authors:** Neal Russell, Wolfgang Stöhr, Nishad Plakkal, Aislinn Cook, James A Berkley, Bethou Adhisivam, Ramesh Agarwal, Manica Balasegaram, Daynia Ballot, Adrie Bekker, Eitan Naaman Berezin, Davide Bilardi, Suppawat Boonkasidecha, Cristina G. Carvalheiro, Suman Chaurasia, Sara Chiurchiu, Simon Cousens, Tim R. Cressey, Tran Minh Dien, Yijun Ding, Angela Dramowski, Madhusudhan DS, Ajay Dudeja, Jinxing Feng, Youri Glupczynski, Herman Goossens, Tatiana Munera Huertas, Mohammad Shahidul Islam, Daniel Jarovsky, Nathalie Khavessian, Meera Khorana, Tomislav Kostyanev, Mattias Larsson, Maia De Luca, Surbhi Malhotra-Kumar, Marisa M. Mussi-Pinhata, Ruchi Nanavati, Sushma Nangia, Jolly Nankunda, Alessandra Nardone, Borna Nyaoke, Christina W Obiero, Maxensia Owor, Wang Ping, Kanchana Preedisripipat, Shamim Qazi, Tanusha Ramdin, Amy Riddell, Emmanuel Roilides, Samir K Saha, Kosmas Sarafidis, Reenu Thomas, Sithembiso Velaphi, Tuba Vilken, Yajuan Wang, Yonghong Yang, Liu Zunjie, Sally Ellis, Julia Bielicki, A Sarah Walker, Paul T. Heath, Mike Sharland

## Abstract

**Background:** Neonatal sepsis is a leading cause of child mortality, and increasing antimicrobial resistance threatens progress towards the Sustainable Development Goals. Evidence to guide antibiotic treatment for sepsis in neonates and young infants from randomized controlled trials or observational studies in low- and middle-income countries (LMICs) is scarce. We aimed to describe patterns of antibiotic use, pathogens and outcomes in LMIC hospital settings globally to inform future clinical trials on the management of neonatal sepsis.

**Methods & Findings:** Hospitalised infants aged <60 days with clinical sepsis were enrolled during 2018-2020 by 19 sites in 11 countries (mainly Asia and Africa). Prospective daily data was collected on clinical signs, supportive care, antibiotic treatment, microbiology and clinical outcome at 28 days. The study was observational, with no changes to routine clinical practice. 3204 infants were enrolled, with median birth weight 2500g (IQR 1400-3000) and postnatal age 5 days (IQR 2-15). Of 309 enrolled aged 28-60 days, 58.6% (n=181) were ex-preterm and/or a neonate at admission. 2215 (69%) infants had been in hospital since birth.

206 different empiric antibiotic combinations were used, which were structured into 5 groups that were developed from the World Health Organisation (WHO) AWaRe classification. 25.9% (n=814) of infants started a WHO first line regimen (Group 1 -Access, penicillin-based regimen) and 13.8% (n=432) started WHO second-line cephalosporins (cefotaxime/ceftriaxone) (Group 2- ‘Low’ Watch). The largest group (34.0%, n=1068) started a regimen providing partial extended-spectrum beta-lactamase (ESBL)/pseudomonal coverage (piperacillin-tazobactam, ceftazidime, or fluoroquinolone-based) (Group 3 – ‘Medium’ Watch), 18.0% (n=566) started a carbapenem (Group 4 – ‘High’ Watch), and 1.8% (n=57) started a Reserve antibiotic (Group 5, largely colistin-based). Predictors of starting non-WHO recommended regimens included lower birth weight, longer in-hospital stay, central vascular catheter use, previous culture positive sepsis or antibiotic exposure, previous surgery and greater sepsis severity. 728/2880 (25.3%) of initial regimens in Group 1-4 were escalated, mainly to carbapenems, and usually for clinical indications (n=480; 65.9%).

564 infants (17.6%) isolated a pathogen from their baseline blood culture, of which 62.9% (n=355) had a Gram-negative organism, predominantly *Klebsiella pneumoniae* (n=132) and *Acinetobacter* spp. (n=72). These leading Gram-negatives were both mostly resistant to WHO-recommended regimens, and also resistant to carbapenems in 32.6% and 71.4% of cases respectively. MRSA accounted for 61.1% of *Staphylococcus aureus* (n=54) isolates.

Overall, 350/3204 infants died (11.3%; 95%CI 10.2-12.5%), with 17.7% case fatality rate among infants with a pathogen in baseline culture (95%CI 14.7-20.1%, n=99/564). Gram-negative infections accounted for 75/99 (75.8%) of pathogen-positive deaths, especially *Klebsiella pneumoniae* (n=28; 28.3%), and *Acinetobacter* spp. (n=24; 24.2%).

**Conclusion:** A very wide range of antibiotic regimens are now used to treat neonatal sepsis globally. There is common use of higher-level Watch antibiotics, frequent early switching and very infrequent de-escalation of therapy. Future hospital based neonatal sepsis trials will ideally need to account for the multiple regimens used as standard of care globally and include both empiric first line regimens and subsequent switching in the trial design.

**Author Summary:** *Why was this study done?:*

➢ Increasing trends in antimicrobial resistance (AMR) disproportionately affect neonates and young infants with sepsis in LMIC settings and undermine the effectiveness of WHO-recommended antibiotics.
➢ Despite this, longitudinal data on antibiotic management strategies and outcomes of affected hospitalised neonates and young infants in LMIC settings are extremely limited, impeding the design of robust antibiotic trials.

*What did the researchers do and find?:*

➢ To our knowledge this is the first global, prospective, hospital-based observational study of clinically diagnosed neonatal sepsis across 4 continents including LMIC settings, with daily data on clinical status, antibiotic use and outcomes.
➢ There was a high mortality among infants with culture positive sepsis (almost 1 in 5), and a significant burden of antibiotic resistance.
➢ This study highlights wide variations in standard of care for sepsis in neonates and young infants with more than 200 different antibiotic combinations, significant divergence from WHO-recommended regimens, and frequent switching of antibiotics.

*What do these findings mean?:*

➢ These data demonstrate that patterns of routine antibiotic use are now markedly divergent from global guidance
➢ There is an urgent need for randomised controlled trials to address optimal empiric first and second line antibiotic treatment strategies in LMIC hospital settings with a significant AMR burden.
➢ Data from this study can inform the design of multicentre hospital-based neonatal antibiotic trials in LMIC settings.
➢ The wide range of multiple antibiotic regimens routinely used as Standard of Care (SOC) suggests the need for novel trial designs.

## Introduction

Sepsis is responsible for a significant burden of disease in neonates and young infants, both as a primary cause of death, and as a frequent contributor.^12^ Access to facility-based delivery and care, including antibiotics, has not consistently reduced mortality to the extent necessary to achieve the Sustainable Development Goal targets in many low and middle income countries (LMICs). ^3^ Antimicrobial resistance (AMR) increasingly threatens to undermine the effectiveness of antibiotics and potentially slows the progress in reducing mortality, particularly in LMICs,^4–10^ with AMR-attributable neonatal deaths recently estimated between 140,000^10^ and 214,000.^11^

Recent large scale antibiotic trials in neonates and young infants in LMIC settings have largely focused on simplification of first line antibiotic regimens with oral amoxicillin and short course gentamicin.^12,13^ These have been based in community settings and included populations with mortality below 2%. However, an increasing global proportion of new-borns are delivered in facilities (∼3/4),^14^ where sepsis case fatality rates and the burden of AMR are greater. Despite this, there is limited high quality evidence generated in LMIC neonatal inpatient settings to guide empiric antibiotic treatment.^4,15^ Published observational data largely involve single centre studies reporting non-systematically collected microbiological data, which are rarely accompanied by detailed clinical and antibiotic use data. Global antibiotic use data largely rely on point prevalence surveys, with limited information on patterns of switching and duration.^16^

In this paper we describe a prospective multi-country observational study in which we collected detailed daily longitudinal data on clinical features, microbiology, antibiotic use and switching, and outcomes of neonatal sepsis in hospital settings, predominantly in LMICs. The primary aim was to describe the variation and patterns of hospital-based antibiotic use to inform the design of hospital based neonatal sepsis antibiotic trials and future guidance.^17^

## Methods

### Study design and participants

Hospitalized infants <60 days of age with a new episode of clinically suspected sepsis were enrolled between 2018 and 2020, in 19 hospital sites across 11 countries in Asia (Bangladesh, China, India, Thailand and Vietnam), Africa (Kenya, South Africa, Uganda), Europe (Italy, Greece) and South America (Brazil). Sites were selected after conducting a feasibility study^18^ to represent diverse regions and to include secondary and tertiary referral hospitals, public facilities, and facilities with varying proportions of in-born and out-born neonates, with access to microbiology.

Infants were eligible if the local physician had decided to treat the infant with antibiotics for a new episode of sepsis meeting the inclusion criteria (supplement figure 1), derived by combining clinical and laboratory criteria from WHO possible Serious Bacterial Infection (pSBI)^19^ and EMA Criteria for neonatal sepsis trials.^20^ To allow for variation in access to laboratory testing, and ensure generalizability to varying LMIC hospital contexts, laboratory values were not mandatory. A minimum of 2 clinical, or 1 clinical and 1 laboratory sepsis criteria, were required for inclusion, and up to 200 infants per site were enrolled according to a sampling frame adapted to local case volume and activity. Infants were excluded if an alternative primary diagnosis other than sepsis was suspected, or a serious non-infective comorbidity was expected to cause death within 72 hours. Previous antibiotic use was not an exclusion criterion as long as a new antibiotic regimen was being started after a blood culture for a distinct new episode of sepsis. Sepsis episodes occurring >48 hours after admission, defined by time of blood culture, were considered healthcare-associated infections (HAI).

Ethical approval was obtained from St. George’s, University of London (SGUL) Research Ethics Committee and sites’ local, central or national ethics committees and other relevant local bodies, where required. Design and reporting were guided by the STROBE-NI framework,^21^ and the trial was registered with ClinicalTrials.gov (NCT03721302).

### Procedures

After consent from parents, baseline demographic and clinical data were collected, followed by prospective daily collection of observational data including multiple clinical parameters, laboratory investigations and microbiological results. Antibiotic data were collected daily including drug, dose, route, duration, switching and reasons given for any changes. Clinical data collection was required up to 48 hours after the completion of antibiotic therapy or discharge if sooner. Aside from a mandatory blood culture at enrolment and daily monitoring of vital signs, all clinical observations and investigations were performed according to routine local site practices. Infants were followed until 28 days after enrolment in-person if still hospitalized, or by telephone post-discharge. A final diagnosis was documented by clinicians, as were primary and secondary causes of death, and any clinical illness or readmission occurring after discharge and within 28 days of enrolment.

Data were collected by research and clinical staff based on clinical observation and routine source documentation (e.g. medical and nursing notes, vital signs and prescription charts), and entered and managed using REDCap™ electronic data capture tools^22^ hosted at SGUL (details on data monitoring in supplementary appendix.)

### Microbiology/Laboratory assessments

Laboratory analysis was performed in each site following local practice, with standard operating procedures developed to optimise procedures including blood culture technique, and antibiotic susceptibility testing. A locally defined algorithm was used to classify contaminants and pathogens by site clinicians and microbiologists. External validation of the capability of laboratories to detect multi-drug resistant (MDR) Gram-negative pathogens from each site was evaluated objectively by testing an external quality assurance (EQA) panel sent from the central laboratory at Laboratory of Medical Microbiology (LMM) at the University of Antwerp (UA).

European Committee on Antimicrobial Susceptibility Testing (EUCAST) or Clinical and Laboratory Standard Institute (CLSI) guidelines and interpretive algorithms were used to interpret reported antibiotic susceptibility testing data for all organisms. EUCAST 2019 breakpoints table (v9.0) and guidelines were used as this was the year the majority of the data was reported. In most cases, where susceptibility to a particular antibiotic was reported, this was used to determine susceptibility. If the organism was considered intrinsically resistant to a particular antibiotic according to EUCAST, then it was coded as resistant regardless of reported susceptibility. If susceptibility to a particular antibiotic was not reported, susceptibility results of a different antibiotic in the same class were used to determine susceptibility (e.g. susceptibility of organism to another aminoglycoside if gentamicin susceptibility was not reported); if no other antibiotic in that class had reported susceptibility then susceptibility was coded as unknown.

### Analysis of antibiotic patterns of use

The initial antibiotic regimen was defined as the first new antibiotic(s) started within 24 hours from baseline blood culture (including 3 hours pre baseline culture). To structure the analysis and reporting of the multiple regimens used, a novel method of grouping antibiotics was derived, based on the Essential Medicine List for Children (EMLc) AWaRe classification (Access, Watch, Reserve; supplementary appendix),^23^ with the ‘Watch’ category divided into 3 distinct groups of ‘Low/Medium/High Watch’ based on inclusion in current WHO guidelines (Low Watch) and likelihood of resistance generation in regimens outside WHO recommendations (Medium or High Watch). ^24^ Antibiotic groups were defined by the main ‘stem’ in the antibiotic combination: Group 1 antibiotics included a first line WHO recommended penicillin-based regimen (e.g. ampicillin and gentamicin) (Access), Group 2 included 3^rd^ generation cephalosporin (eg cefotaxime/ceftriaxone)-based WHO regimens (‘Low’ Watch), Group 3 included regimens with partial anti-extended-spectrum beta-lactamase ((ESBL) or pseudomonal activity (e.g. piperacillin-tazobactam/ceftazidime/fluoroquinolone-based) (‘Medium’ Watch), and Group 4 included carbapenems (e.g. meropenem) (‘High’ Watch). Group 5 antibiotics included Reserve antibiotics targeting carbapenem resistant organisms (e.g. colistin). Aminoglycosides (e.g. gentamicin/amikacin), glycopeptides (e.g. vancomycin/teicoplanin) and metronidazole used in combination regimens were classified as additional coverage and did not define the main antibiotic ‘stem’ for the grouping. All antifungals and antivirals were excluded from the antimicrobial treatment data as these were not relevant to the analysis. Escalation of treatment was defined as a switch to a higher group antibiotic, and de-escalation was defined as switching to a lower group or discontinuation of the ‘stem’ antibiotic whilst continuing with an additional coverage antibiotic.

### Statistical analysis

The pre-specified primary outcome was mortality through 28 days post-enrolment, analysed using Kaplan-Meier and Cox proportional hazards regression with site-level random effects. Time was measured from the initial blood culture sample, censoring at the earliest of day 28, withdrawal or last contact if lost post-discharge. To analyse whether time from baseline culture to the start of new antibiotic regimen or type of pathogen were associated with mortality, respectively, we adjusted for previously established baseline predictors of mortality (Submitted manuscript^25^). Multivariable logistic regression with backwards elimination (exit p=0.05) adjusted for site was used to analyse factors associated with starting group 3-5 versus group 1-2 regimens in sites with at least 10% of infants in either. Candidate factors were known at sepsis presentation and included birth weight, gestational age, postnatal age, time in hospital, central line or indwelling catheter, Intravenous (IV) antimicrobials in previous 24 hours, signs of meningitis, previous positive culture, previous surgery, and a score reflecting sepsis severity (Submitted manuscript^25^). Cumulative incidence antibiotic escalation or stop of all iv antibiotics was estimated with death as competing risk.

Analyses used Stata version 16.1.

### Role of the funding source

This study was funded by the Global Antibiotic Research and Development Partnership (GARDP), made possible by Bill & Melinda Gates Foundation; German Federal Ministry of Education and Research; German Federal Ministry of Health; Government of the Principality of Monaco; the Indian Council for Medical Research; Japanese Ministry of Health, Labour and Welfare; Netherlands Ministry of Health, Welfare and Sport; South African Medical Research Council; UK Department of Health and Social Care (UK National Institute of Health Research and the Global Antimicrobial Resistance Innovation Fund – GAMRIF); UK Medical research Council; Wellcome Trust. GARDP has also received core funding from the Leo Model Foundation; Luxembourg Ministry of Development Cooperation and Humanitarian Aid; Luxembourg Ministry of Health; Médecins Sans Frontières; Swiss Federal Office of Public Health; UK Foreign, Commonwealth & Development Office (previously the UK Department for International Development).

The authors had access to all study-related data and had final responsibility for publication.

## Results

3204 infants (90.4% neonates aged <28 days, n=2895; 42.1% female, n=1348) were recruited from 20^th^ August 2018 - 29^th^ February 2020 (table 1; supplement figure 2). Sites included varying populations of infants, and levels of care (see supplement figures 3-5). The median postnatal age was 5 (IQR 1-15) days, and 3088 (96.4%) infants had been born in a hospital/facility (1550 in the enrolling facility), 1412 (44.3%) by caesarean section (969 as an emergency). 71 (2.2%) had previously been treated for an episode of culture-positive sepsis. The median (IQR) gestational age at birth was 37 (31-39) weeks, with birth weight 2500g (1400-3000g). At enrolment, 69.1% (n=2215) infants had been hospitalised since birth, and 30.9% (n=989) were admitted from the community. 2759 (86.1%) were recruited in a neonatal unit. Among 309 (9.6%) infants enrolled aged ≥28 days, the majority (n=181; 58.6%) were either ex-premature (n=146; 47.4%) and/or had been admitted during the neonatal period (n=136, 44.2%).

**Table 1:**
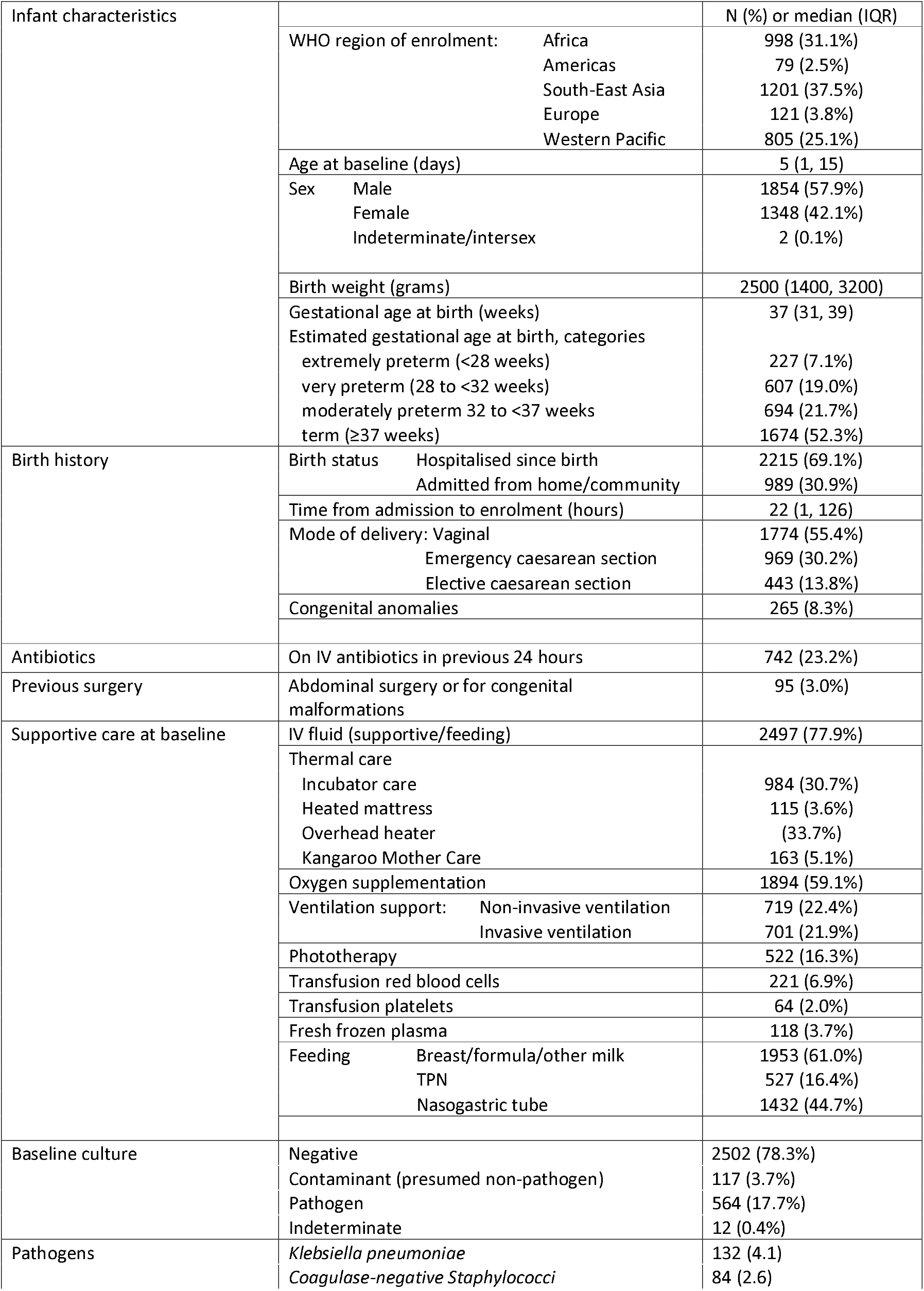
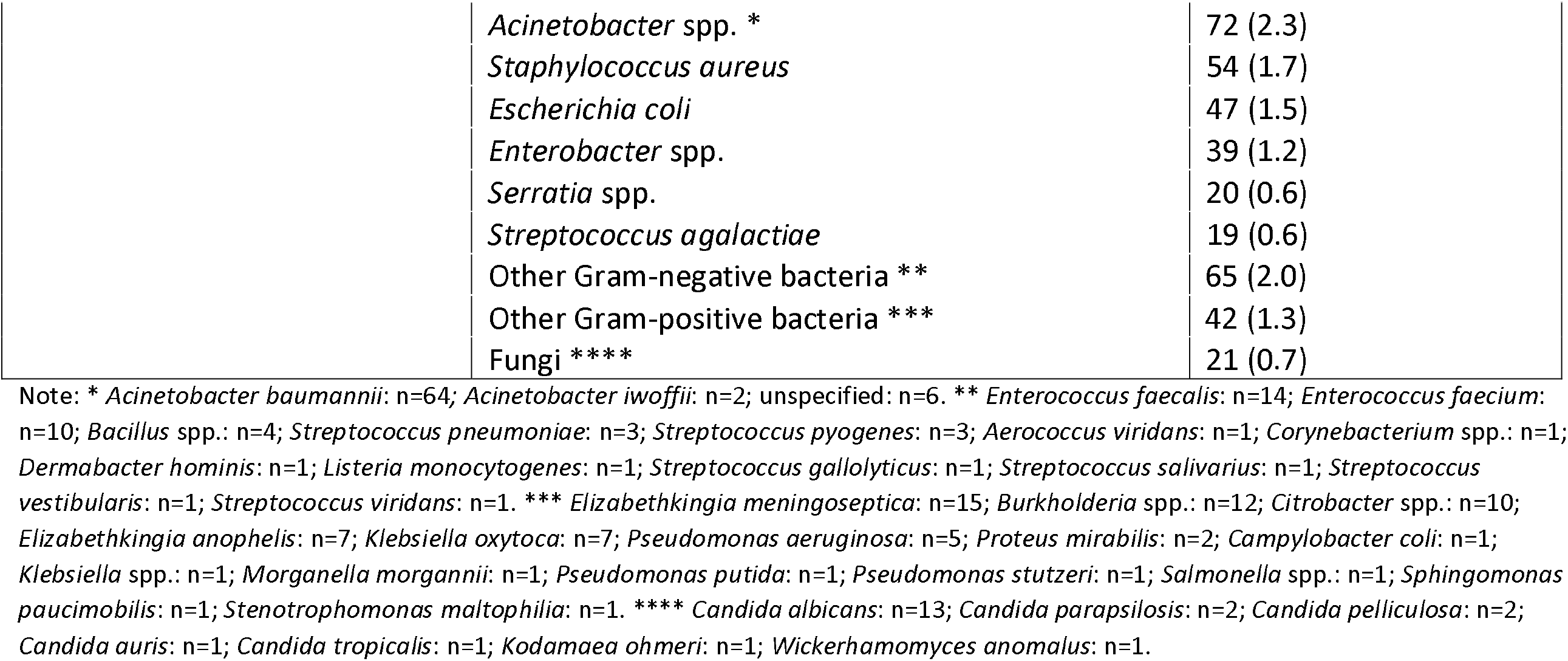
Baseline Characteristics and Antibiotic use.

The most common previously identified risk factors for sepsis other than prematurity were preterm premature rupture of membranes (14.5%, n=466), prolonged rupture of membranes (>18hrs) (10.2% n=328), prelabour rupture of membranes at term (9.4%, n=300), presence of an indwelling central vascular catheter (8.2% n=262), intrapartum fever >38°C (3.5%, n=112), chorioamnionitis (2.8%, n=75), known maternal Group B streptococcus (GBS) colonisation (1.4%, n=46) and previous surgery (abdominal or for congenital malformations: 3.0%, n=95). Of note, 8.3% (n=265) had a congenital anomaly (supplement table 1), and 7% (n=220) of infants were exposed to maternal HIV. 1318 (41.1%) sepsis episodes were healthcare-associated (occurring >48h after hospital admission).

### Clinical and laboratory findings

A median of 4 (IQR 2-5) clinical signs were present at baseline, the most common being respiratory (65.8%, n=2107), difficulty feeding (45.7%, n=1464), lethargy or reduced movement (35.3%, n=1,131), abdominal distension (24.3%, n=777) and evidence of shock (21.3%, n=683). Signs suggestive of meningitis were reported in <10% of babies (irritability, convulsions, abnormal posturing, bulging fontanelle) (supplement figures 6-7). The availability of laboratory values varied by site. At baseline, a base excess <-10 mmol/L was documented in 352 of 1492 with results available (23.6%), lactate >2mmol in 1034/1283 (80.6%), raised CRP >10mg/L in 1306/2286 (57.1%), abnormal white blood cell count (<4 or >20×10^9^ cells/L) in 875/2800 (31.3%), and thrombocytopenia (<150×10^9^ cells/L) in 619/2776 (22.3%) (supplement table 2).

### Patterns of empiric antibiotic use

1180 (36.8%) infants had a history of previous intravenous antibiotic treatment, and 742 (23.2%) had been receiving an intravenous antibiotic in the previous 24h before starting new antibiotics for the new episode of sepsis. Median time from baseline culture being taken to new intravenous antibiotic treatment being started for the distinct sepsis episode was 1 hour (IQR 0-3); 2913 (90.9%) started within 8 hours, and 228 (7.1%) within 8-24 hours (supplement figure 8).

There were 206 different combinations of empiric antibiotics started at baseline in this hospital-based cohort. These were grouped as described in the methods, and most frequent regimens are reported in table 2. 25.9% (n=814) of infants were started on a WHO-recommended first line penicillin-based regimen (Group 1 - Access). 13.8% (n=432) of infants were started on a WHO second line cefotaxime or ceftriaxone-based combination (Group 2 – ‘Low’ Watch) (figure 1a). The largest group (34.0%, n=1068) were started on a regimen providing partial ESBL/pseudomonal coverage (piperacillin-tazobactam, ceftazidime, or fluoroquinolone based) (Group 3 – ‘Medium’ Watch). Within Group 3 ceftazidime ± amikacin (n=436; 13.9%) and piperacillin/tazobactam±amikacin (n=410; 13.1%) were prominent combinations. 18.0% (n=566) of initial regimens were carbapenem-based (Group 4 – ‘High’ Watch), among whom meropenem ± vancomycin (n=447; 14.2%) was the most common combination. 1.8% (n=57) of initial regimens were classified as group 5 antibiotics targeting carbapenem-resistant organisms, predominantly colistin-based (Group 5 - Reserve) (table 2; supplement figures 9-13). An ‘Other’ group (n=204) consisted of more rarely used local regimens not on the WHO EMLc, or regimens which did not include a new antibiotic ‘stem’ that was used to define groups 1-5 (e.g. aminoglycoside or glycopeptide given alone or in combination with each other) (supplementary figure 14). Cefoperazone/sulbactam (n=99) was the most common antibiotic in this category, but was used as initial regimen only in India (n=86; 14.5%), China (n=11; 1.9%) and Vietnam (n=2; 1.0%).

**Table 2:**
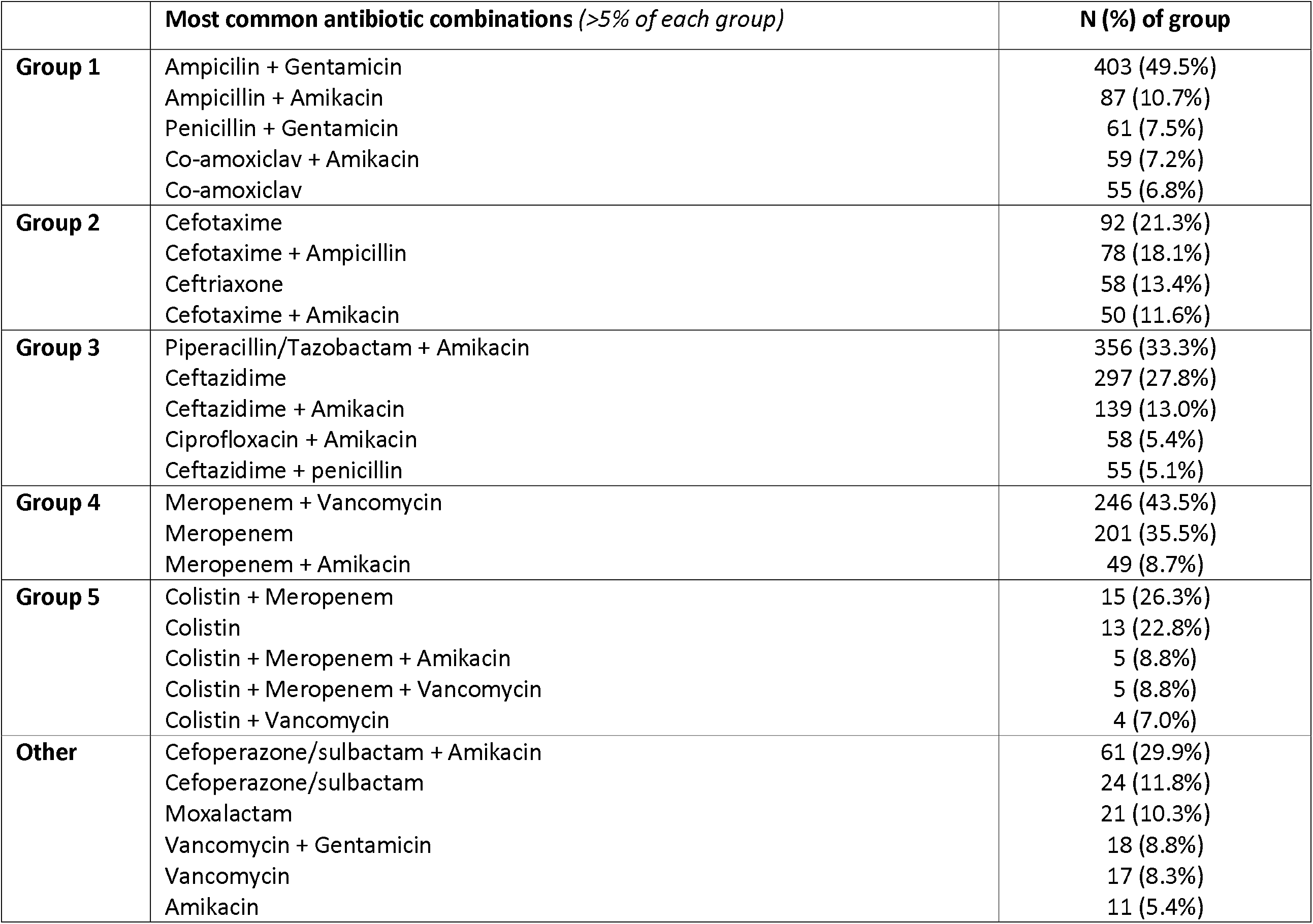
Most common antibiotic regimens in each Group.

**Figure 1.**
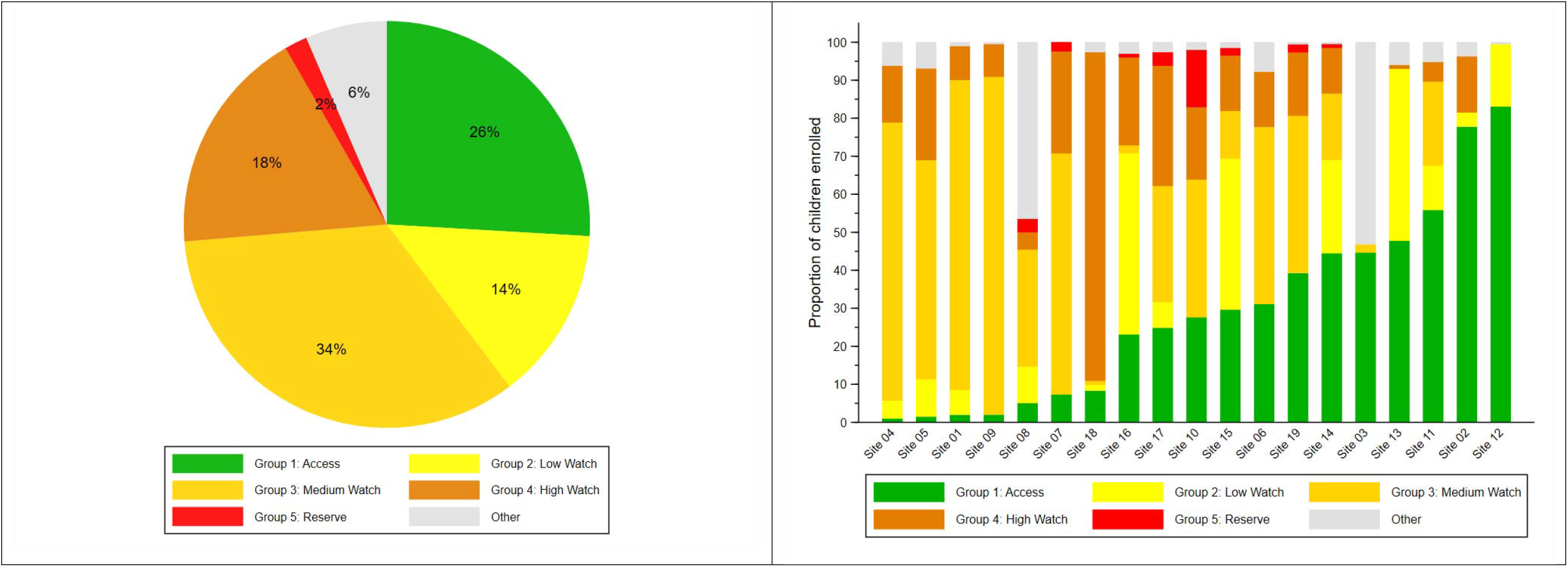
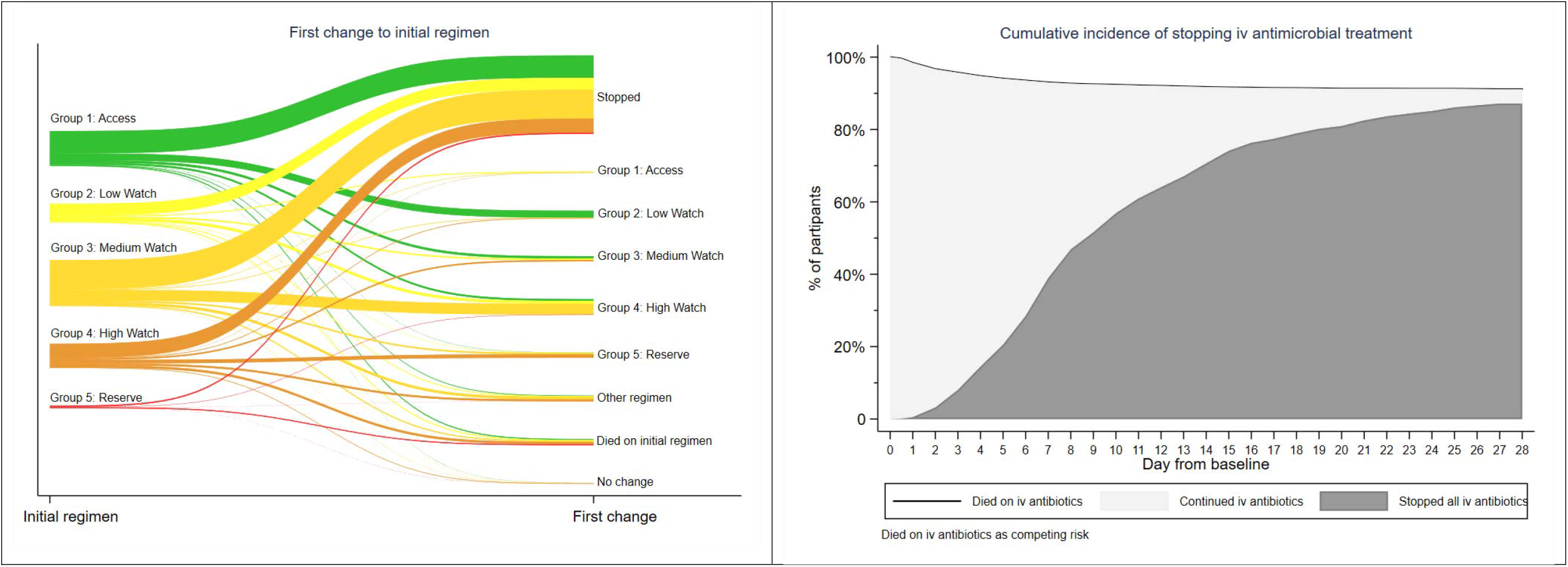
1a Antibiotic AwaRe Wheel including empiric antibiotic grouping at baseline; 1b Empiric baseline therapy by site; 1 c First change of initial regimen, by baseline regimen; 1d Cumulative incidence of stopping all iv antibiotics. Group 1 = First line WHO recommended penicillin-based regimen (e.g. ampicillin and gentamicin) (Access). Group 2 = 3^rd^ generation cephalosporin (eg cefotaxime/ceftriaxone)-based WHO regimens (‘Low’ Watch). Group 3 = regimens with partial anti-extended-spectrum beta-lactamase ((ESBL) or pseudomonal activity (e.g. piperacillin-tazobactam/ceftazidime/fluoroquinolone-based) (‘Medium’ Watch). Group 4 = Carbapenems (‘High’ Watch). Group 5 = Reserve antibiotics targeting carbapenem resistant organisms (e.g. colistin).

There was wide variation between sites in the frequency of empiric use of different antibiotic groups (Figure 1b). Some sites used predominantly Group 1 antibiotics as the initial regimen, and others often started immediately with Group 3 or 4 regimens, or a mixture of all groups. The most frequently prescribed empiric regimens used for healthcare associated infections (HAI) were meropenem±vancomycin (28.2%, 361/1280), followed by piperacillin/tazobactam±amikacin (17.5%, n=224), ceftazidime±amikacin (6.7%, n=86, colistin (4.3%, n=55) and cefoperazone/sulbactam (3.9%, n=50). Ampicillin+gentamicin was the most common regimen for non-HAI (19.7%, 366/1861), followed by ceftazidime±amikacin (18.8%, n=350) and piperacillin/tazobactam±amikacin (10.0%, n=186) (supplement table 3).

Adjusting for site, predictors of starting empiric therapy with group 3-5 rather than group 1-2 antibiotics included lower birth weight (OR=0.57 per additional kg, 95% CI 0.47-0.69), presence of a central vascular catheter (OR=3.48, 95% CI 1.74-6.94), previous antibiotics at enrolment (OR=5.71, 95% CI 3.73-8.77), previous culture positive sepsis (OR=25.71, 95% CI 3.00-220.7), longer time in hospital (48 versus 0 hours: OR=4.41, 95% CI 3.41-5.69), previous surgery (OR=5.18, 95% CI 1.65-16.28) and higher sepsis severity (OR=1.27 per additional score point, 95% CI 1.16-1.40) (see supplement table 4).

### Antibiotic switching

After initial therapy, 728/2880 (25.3%) who started on Group 1-4 regimens were escalated to a higher group regimen, the majority switching within the first days of treatment (supplement figures 15-17). 258/814 (31.7%) infants escalated from Group 1 (ampicillin/gentamicin) regimens, the majority of which switched to Group 2 (cefotaxime/ceftriaxone-based) regimens (61.6%, n=159) (figure 1c). 101/432 (23.4%) infants escalated from Group 2 regimens, mostly directly to a Group 4 carbapenem-based regimen (62.4%, n=63) rather than a Group 3 partial ESBL/pseudomonal activity regimen (32.7%, n=33). 287/1068 (26.9%) escalated from group 3 regimens, and of 566 infants starting treatment with carbapenems (Group 4), 82 (14.5%) escalated therapy to a colistin-containing regimen. Common reported reasons for first escalation of antibiotics overall included clinical deterioration (65.9%, n=480), microbiology results (15.4%, n=112), and worsening inflammatory biomarkers (9.8%, n=71). De-escalation of antibiotics was rare (173/2937; 5.9%).

Cumulative incidence of stopping intravenous treatment within 7 days from baseline culture was 38.9% (95% CI 37.2-40.6%) overall (Figure 1d); 45.8% (95% CI 43.8-47.6%) in pathogen-negative and 6.9% (95% CI 5.0-9.2%) in pathogen-positive cases. After stopping intravenous antibiotics, 350/2803 (12.5%) switched to oral therapy. 289 of 2803 infants who had stopped (10.3%), restarted intravenous antibiotics during the original hospital stay, and a further 84 infants after discharge. 115 (3.4%) were still on uninterrupted antibiotic treatment at day 28. Median total number of days on intravenous antibiotics during the 28 days follow-up was 8 (IQR 6-14) days (supplement figure 18). Of note, intramuscular use of antibiotics was very rare (<0.1%).

### Microbiology

Initial blood culture results were available for 3195 (99.7%) infants; 693 (21.7%) grew at least one organism. Organisms identified as significant pathogens were isolated in 564/693 (>1 pathogen in 29) blood cultures, contaminants (presumed non-pathogens) in 117, and indeterminate in 12 cultures (table 1). Gram-negative and Gram-positive pathogens were found in 62.9% (355/564) and 34.8% (196/564) of infants, respectively (n=8 with both), and fungal pathogens in 21. Amongst infants with a significant pathogen, *Klebsiella pneumoniae* (23.4%, n=132), Coagulase-negative staphylococci (CoNS) (14.9%, n=84), *Acinetobacter species* (12.8%, n=72), *Staphylococcus aureus* (9.6%, n=54), and *Escherichia coli* (8.3%, n=47) were the most common (table 1; supplement figure 19) but with differences between sites (supplement figure 20). *Streptococcus agalactiae* was found in only 19 babies (3.4%). All the common pathogens were identified in both early and late onset sepsis (figure 2), although *Escherichia coli* was more common in the first 3 days of life while *Klebsiella pneumoniae* more frequent in late onset sepsis (supplement table 5).

**Figure 2:**
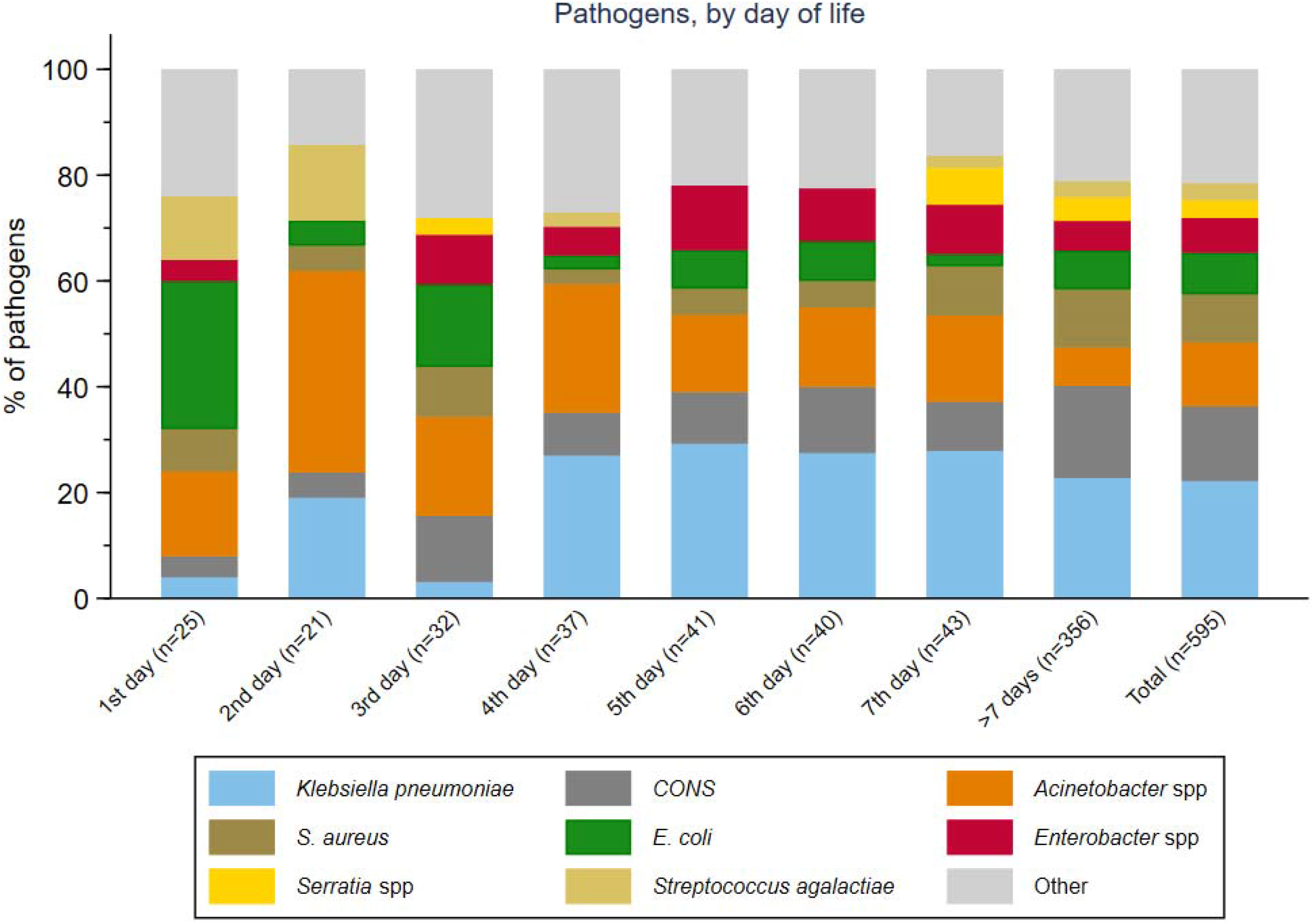
Pathogens isolated in baseline blood culture, by day of life.

58% (75/130 tested) of *Klebsiella pneumoniae* isolates were resistant to gentamicin, 75.0% (96/128) to commonly used 3^rd^ -generation cephalosporins (cefotaxime/ceftriaxone), 46.5% (53/114) were resistant to piperacillin-tazobactam, 46.6% (48/103) to ciprofloxacin, and 32.6% (43/132) to meropenem (supplementary table 6). *Acinetobacter species* were resistant to meropenem in 71.4% (50/70) of cases. *Escherichia coli* retained greater susceptibility to 3^rd^ -generation cephalosporins (64.4%, 29/45 susceptible), and was susceptible in 90.4% (38/42) of cases to piperacillin-tazobactam. Among Gram-negatives there were important differences in susceptibility among aminoglycosides, with amikacin providing significantly better activity than gentamicin (e.g. 61.5% vs 38.5% susceptibility among *Klebsiella pneumoniae*).

Among 54 *Staphylococcus aureus* isolates, 33 (61.1%) were methicillin resistant. *Staphylococcus aureus* and CoNS were susceptible to vancomycin in all of 42 and 73 isolates tested respectively, and all 19 *Streptococcus agalactiae* isolates were susceptible to ampicillin. Other rarer and more site-specific pathogens with high rates of resistance to antibiotics included *Serratia species, Burkholderia species*, and *Elizabethkingia meningoseptica* (supplement table 6).

1226/3204 (38.3%) infants had a cerebrospinal fluid (CSF) culture performed at baseline or during the subsequent 7 days. 73/1226 (6.0%) were culture-positive, 47 with a pathogen, and 26 contaminant/indeterminate. Gram-negative organisms also dominated in CSF cultures (supplement table 7).

### Mortality

Overall, 350 infants (11.3%; 95%CI 10.2-12.5%) died within 28 days of baseline blood culture. Mortality among infants with a pathogen-positive baseline culture was 17.7% (99/564; 95%CI 14.7-21.1%) compared with 9.9% (250/2631; 95%CI 8.8-11.2%) in infants without pathogens (p<0.001). Mortality was higher in infants with Gram-negative (21.3%; 95%CI 17.4-25.9%) or fungal pathogens (38.1%; 95%CI 21.2-61.9%) than with Gram-positive pathogens (8.5%; 95%CI 5.3-13.5%; p<0.001). Mortality was 33.3% (95%CI 23.7-45.5%) in infants with *Acinetobacter* spp., 21.5% (95%CI 15.4-29.6%) with *Klebsiella pneumoniae*, 21.1% (95%CI 8.5-46.8%) with *Streptococcus agalactiae*, 12.8% (95%CI 6.0-26.3%) with E. coli, 11.1% (95%CI 5.2-23.1%) with *Staphylococcus aureus*, and 3.6% (95%CI 1.2-10.8%) with CoNS (figure 3; supplement table 8). Overall, Gram-negative infections accounted for 75/99 (75.8%) of culture-positive deaths, especially *Klebsiella pneumoniae* (n=28; 28.3%), and *Acinetobacter* spp. (n=24; 24.2%). There was significant variation in mortality between different sites (Figure 5).

**Figure 3:**
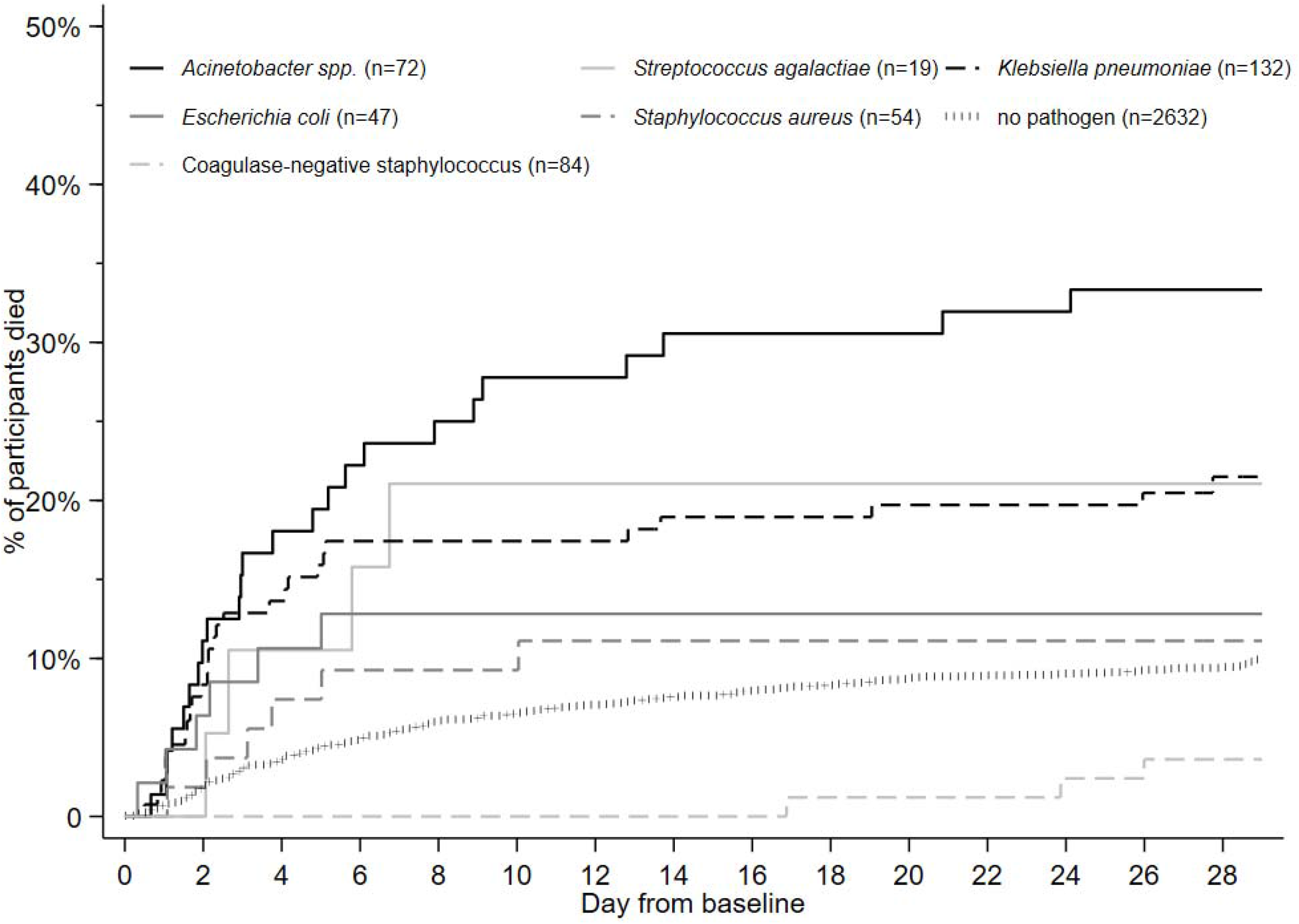
Mortality by pathogen in baseline blood culture. Note: Kaplan-Meier analysis, unadjusted.

**Figure 4:**
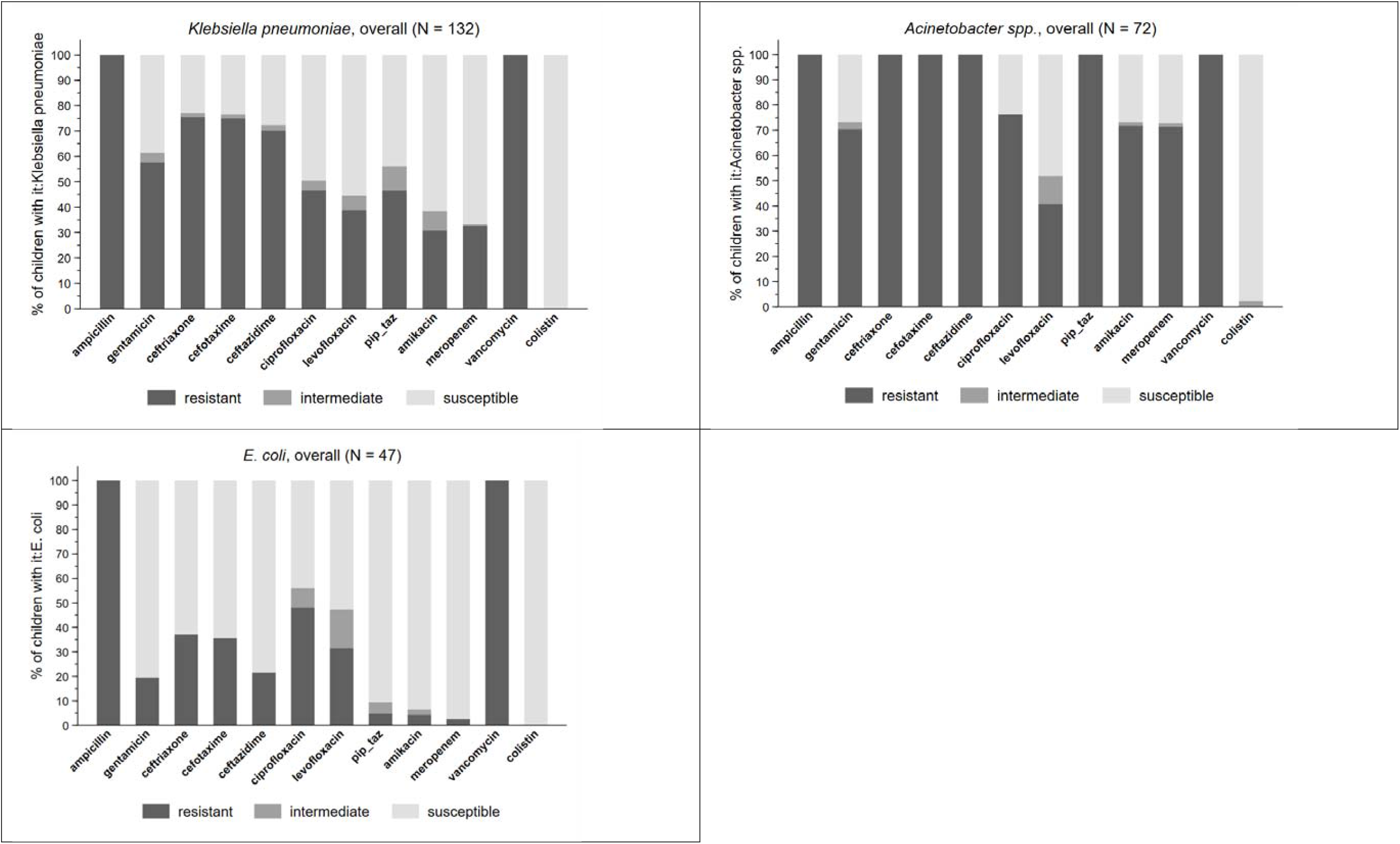
Antibiotic resistance among leading pathogens in baseline blood culture.

**Figure 5:**
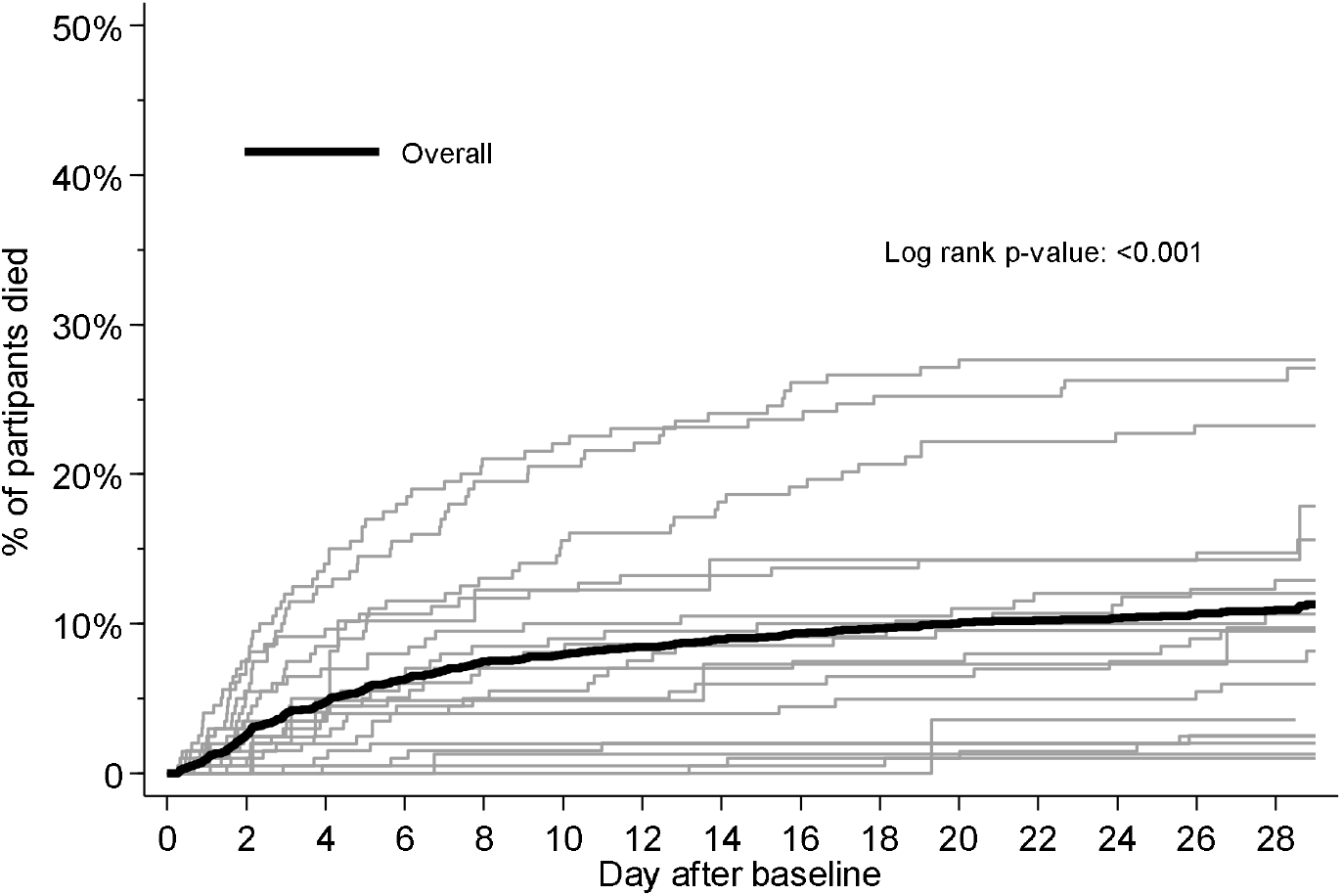
Kaplan-Meier curves for mortality, overall and by site.

There was no significant association between time from blood culture to start of new antibiotics and mortality, neither overall (HR=1.01 per additional 2 hours [95%CI 0.96-1.07]; p=0.68) nor in the subgroup with no previous antibiotic exposure (HR=1.02 [95%CI 0.94-1.10]; p=0.67) (supplement figure 21).

## Discussion

In this hospital-based observational study in 11 countries across Africa, Asia, Europe and Latin America we observed wide variation in antibiotic prescribing beyond WHO recommendations for neonates and young infants with sepsis, with over 200 different empiric combinations, frequent empiric escalation and rare de-escalation of therapy. Antimicrobial resistance was common, and Gram-negative pathogens (*Klebsiella* and *Acinetobacter*), were largely resistant to WHO recommended regimens. Among Gram positives, MRSA accounted for over half of *Staphylococcus aureus* isolates. 10% of pathogen-negative, and 18% of pathogen-positive infants died, and Gram-negative infections were associated with the majority of pathogen-positive deaths.

To our knowledge this is the largest multi-country hospital based observational study of sepsis in neonates and young infants to collect detailed daily prospective clinical and antimicrobial prescribing data linked to microbiology and clinical outcomes. These data confirm trends in antibiotic use reported in recent point prevalence surveys in LMIC settings,^16^ but this study provides evidence on patterns of switching and escalation of therapy. In particular, a third of infants that had started on WHO recommended regimens escalated to broader spectrum antibiotic regimens, and escalation overall was most commonly to a carbapenem. There was also considerable use of ‘carbapenem-sparing’ regimens with ‘partial’ ESBL and anti-pseudomonal activity that varied between centres (piperacillin-tazobactam, ceftazidime and quinolone-based regimens, often in combination with amikacin). Evidence on carbapenem-sparing regimens is limited in neonates,^26^ but *Pseudomonas* infections were rare in NeoOBS and in the recent published literature.^5,6^ Notably, a small but important proportion of infants in some sites received treatment for proven or suspected carbapenem-resistant infection with colistin, for which a recommended approach to dosing and combination therapy remains unclear, and CSF penetration and side effect profile are sub-optimal. ^27^ Some countries are also using less widely used combinations such as cefoperazone/sulbactam, for which neonatal data are limited.^28,29^

The extent to which increasing prevalence of AMR is associated with excess mortality, and whether this may be modifiable with different antibiotic treatment strategies, is unclear in neonates and young infants. AMR has been demonstrated as an independent predictor of mortality in adults with bloodstream infections, including in LMIC settings.^30^ However, data in neonates and young infants in many LMIC settings are scarce.^15,31^ A limited number of largely retrospective observational studies have shown an increase in mortality particularly in association with resistant Gram negative infections^32–35^, such as those due to organisms producing extended spectrum beta-lactamases (ESBL)^3 -39^ and carbapenem resistant organisms (CRO),^40–43^ and some suggest an increase in mortality in the absence of appropriate antimicrobial therapy.^32,44–47^ A recent large multicentre neonatal sepsis study (BARNARDS) demonstrated high resistance to ampicillin + gentamicin (97% and 70%, respectively) among Gram-negative infections; however, analysis of the influence of antibiotic treatment on outcomes was confounded by country effects, and limited clinical data prevented adjusting for other important confounders.^9^ Appropriate analyses of the causal relationship between discordant antibiotic treatment and mortality would need to consider baseline confounders as well as time-dependent confounding,^48^ but to our knowledge this has not been done for neonatal sepsis in LMIC settings to an extent that it could inform changes to global guidance.^9,35,42,49,50^ Relevant analyses in the NeoOBS cohort are ongoing.

Limitations should be taken into account when considering the generalisability of these data. Most sites in NeoOBS represented secondary or tertiary hospitals in urban settings, in some cases receiving referrals after prior treatment in other hospitals. In many cases these facilities provided a higher level of care than is typical in many low resource settings. AMR may be more important, and a wider range of antibiotics may be available in these settings compared to more rural low resource settings.^51,52^ This is an important bias in most studies on AMR in low resource settings, where the need for high quality microbiology means certain settings are over-represented. A significant proportion (36.8%) of infants had previously received antibiotics before the new sepsis episode, which may influence culture yield and pathogen characteristics. Selection bias was also possible beyond site selection, in particular with prospective recruitment potentially leading to a milder phenotype as infants who die rapidly with sepsis are more difficult to enrol, possibly leading to underestimation of mortality. The sites were selected to be heterogenous, with wide variation in levels of supportive care and mortality. These data highlight the major limitations of basing antibiotic recommendations on observational data alone. Analyses to determine the impact of antibiotic use on outcomes are confronted with important biases,^48^ including the influence of initial sepsis severity on antibiotic choice and timing of first administration, antibiotic availability and affordability, inter-site population heterogeneity, local guidelines and microbiology, and varying levels of supportive care.

In the context of increasing resistance to WHO recommended therapy for neonatal sepsis in LMIC settings, and a lack of evidence to guide optimal management due to the limitations of observational data, further randomised antibiotic trials are urgently needed. There is a very limited pipeline of new antibiotics for Gram negative infections^53^ and future strategic trials of novel regimens need to include older off-patent antibiotics.^54,55^ This study demonstrates there is no single “standard of care” in neonatal sepsis. Novel trial designs are therefore needed to define optimal treatment from a background of wide variation in empiric antibiotic prescribing and frequent switching. This study has now informed the design of the NeoSep1 trial (ISRCTN 48721236) which will use a network meta-analytic approach to rank novel off patent antibiotic combinations compared to WHO recommended and other commonly used regimens, combined with a SMART (Sequential Multiple Assessment Randomised Trial) design to allow randomisation to both empiric first and second line treatment.

## Data Availability

Any data request should be made to GARDP initially.

