## Supplementary appendix for "Patterns of antibiotic use, pathogens and clinical outcomes in hospitalised neonates and young infants with sepsis in the NeoOBS global neonatal sepsis observational cohort study"

1. **Supplement figure 1: Clinical and laboratory sepsis enrolment criteria**

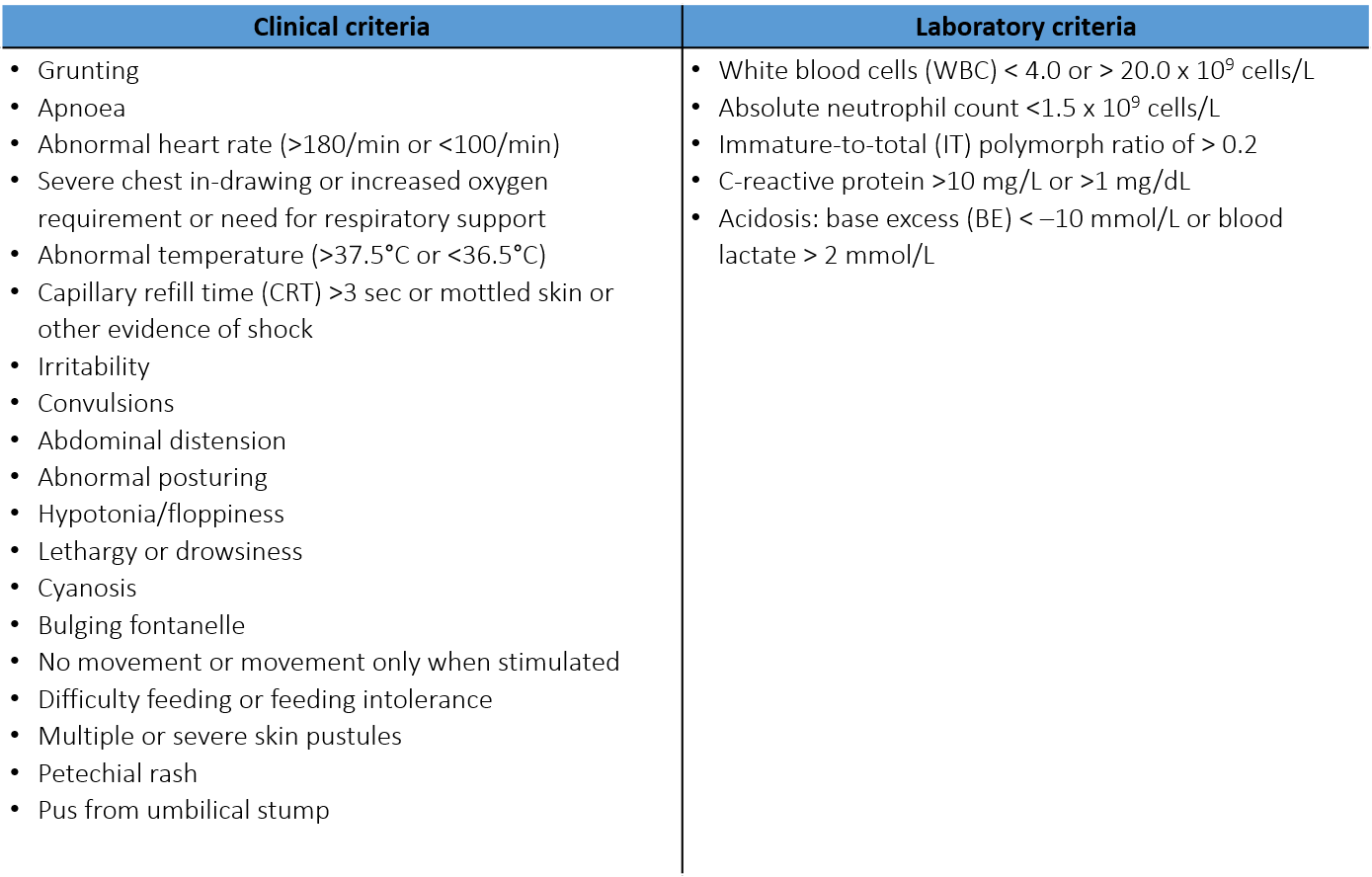

To be identified with **significant sepsis**, infants must meet at least **TWO** criteria, **ONE** of which must be clinical

2. **Supplement figure 2: Flow chart**

Enrolled: 3204

Withdrawn: 5

Died in initial hospital: 315 (10%)

Transferred: 66 (2%)

Alive at day 28: 61 Died: 3 Lost: 2

Discharged: 2011 (63%)

Alive at day 28: 1924 Died: 32 Lost: 55

Hospitalised to day 28: 807 (25%)

3. **Supplement figure 3: Birth weight by site**

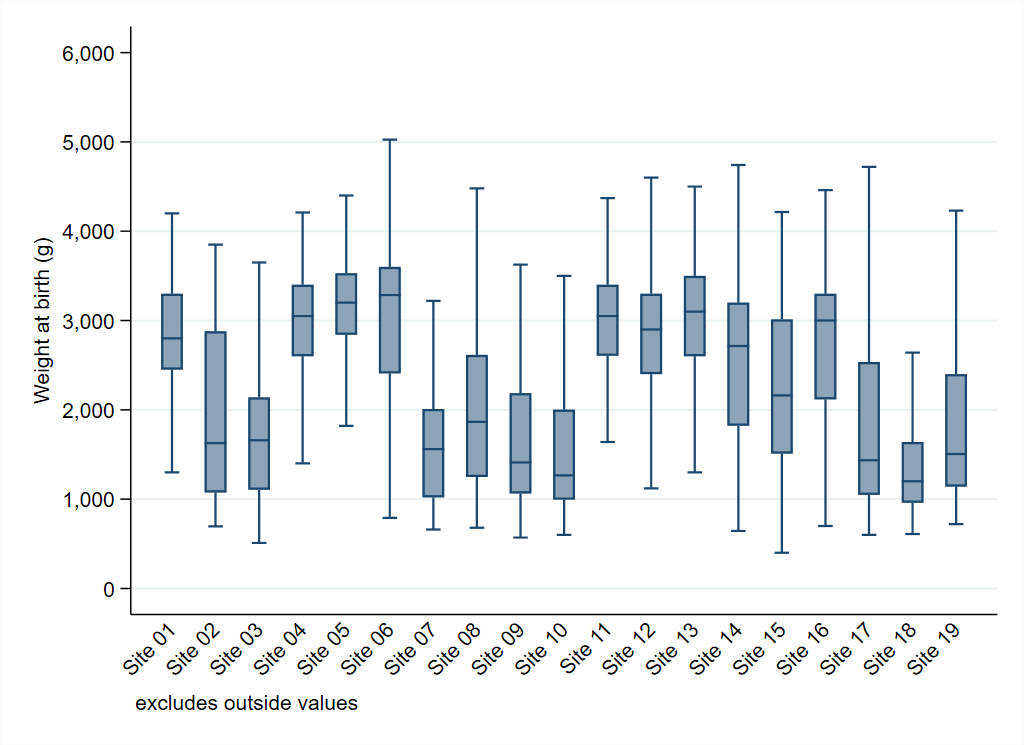

4. **Supplement figure 4: Hospitalised since birth vs admitted since birth, by site**

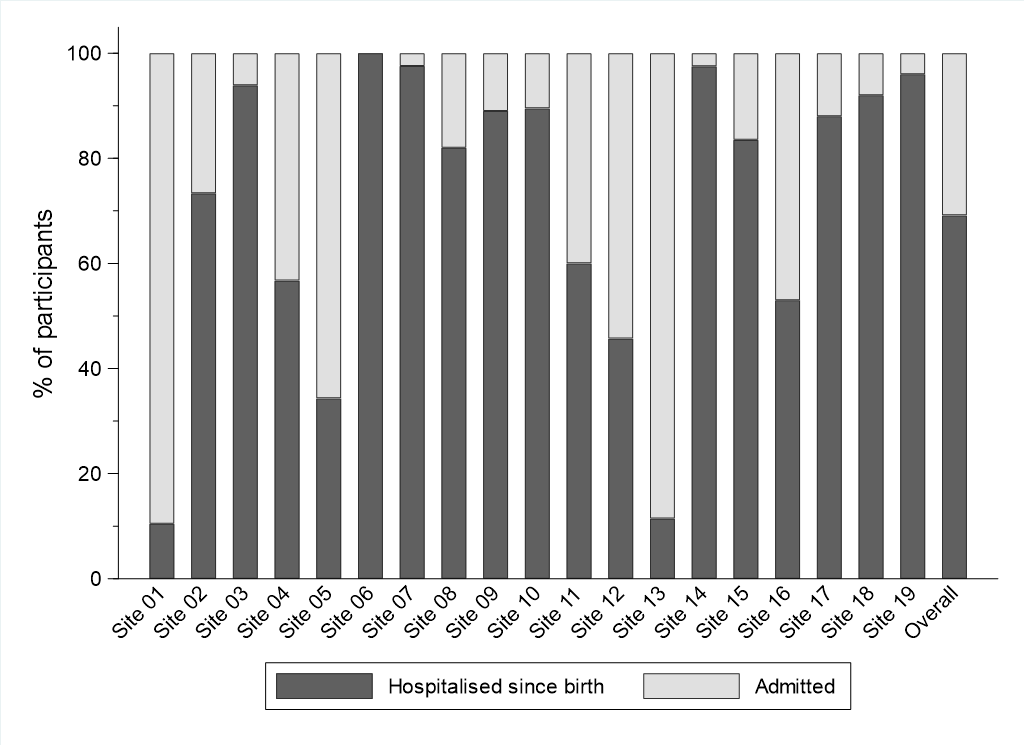

5. **Supplement figure 5: Postnatal age at enrolment by site**

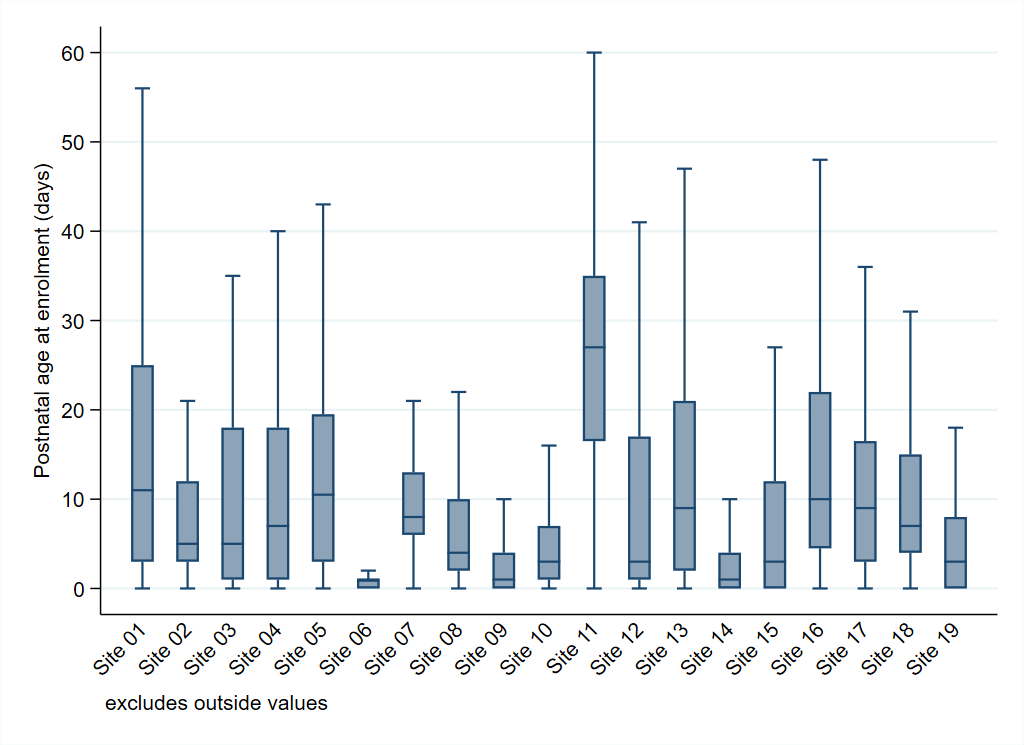

6. **Supplement figure 6: Clinical signs at enrolment (>10%)**

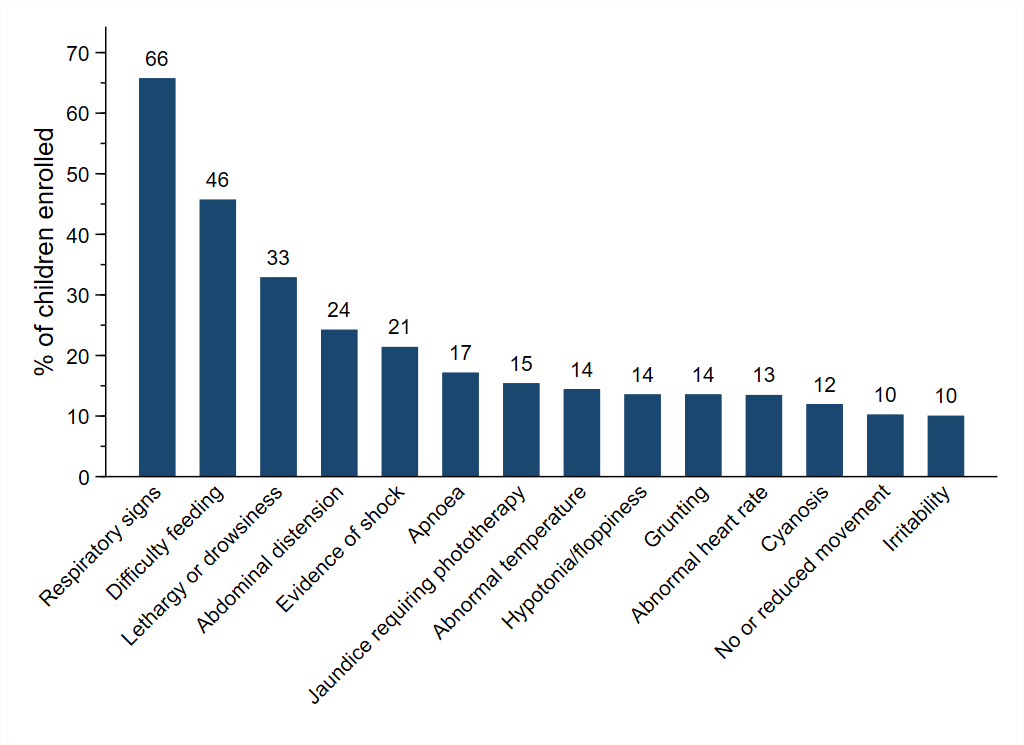

7. **Supplement figure 7: Clinical signs at enrolment (≤10%)**

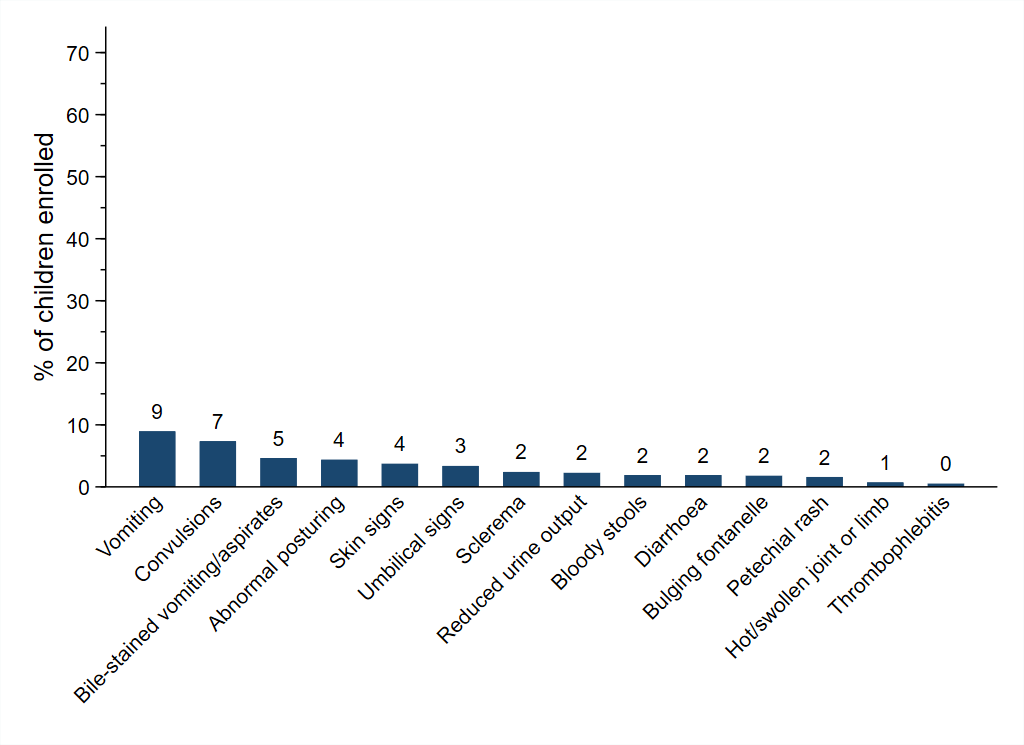

8. **Supplement figure 8: Time to start IV antibiotics after baseline culture, by previous antibiotic exposure**

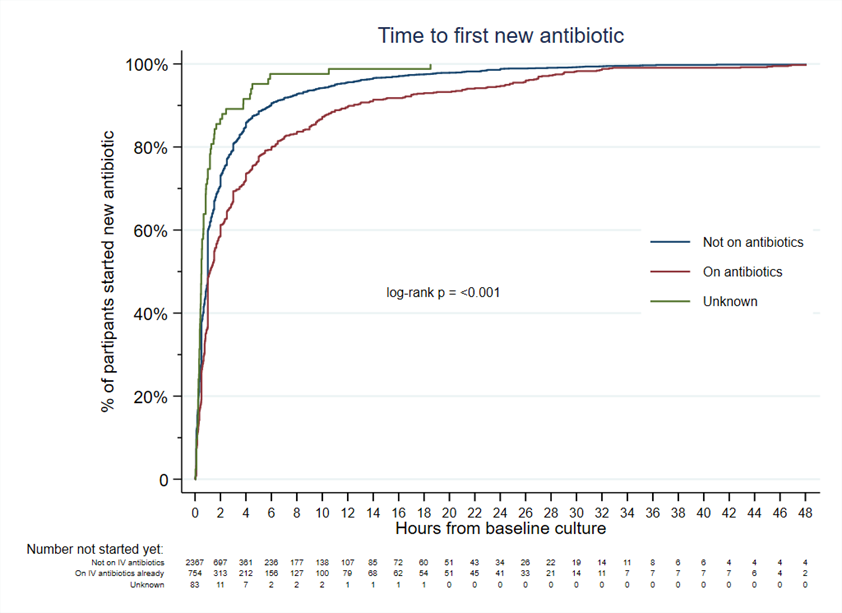

9. **Supplement figure 9: Group 1 Antibiotics - Common combinations**

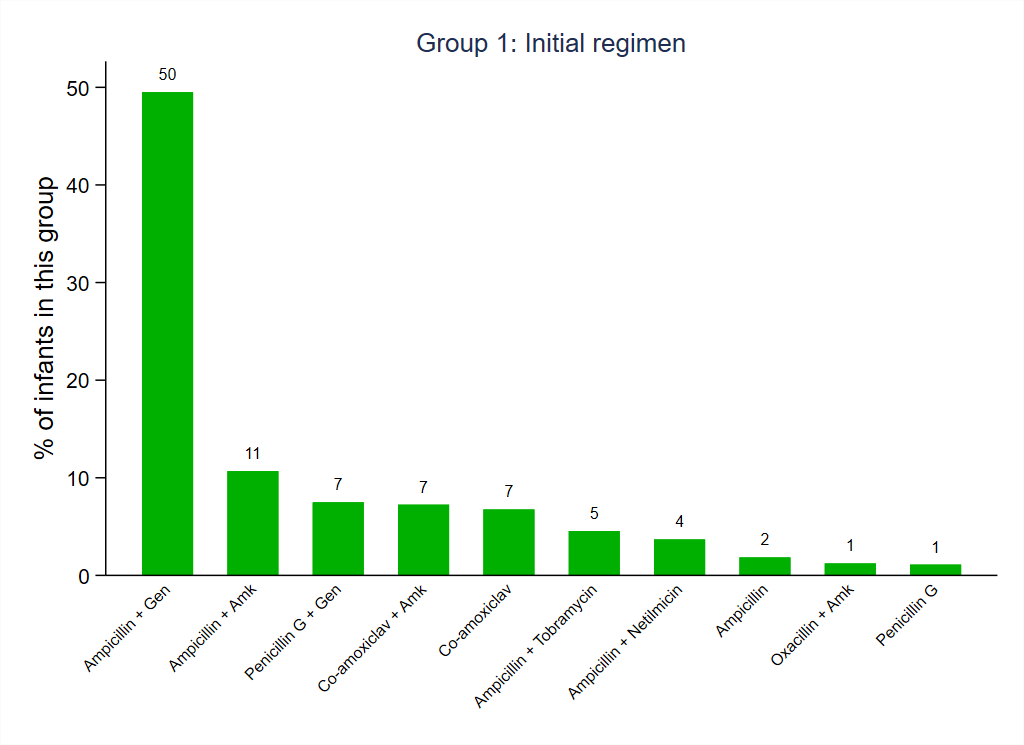

10. **Supplement figure 10: Group 2 Antibiotics - Common combinations**

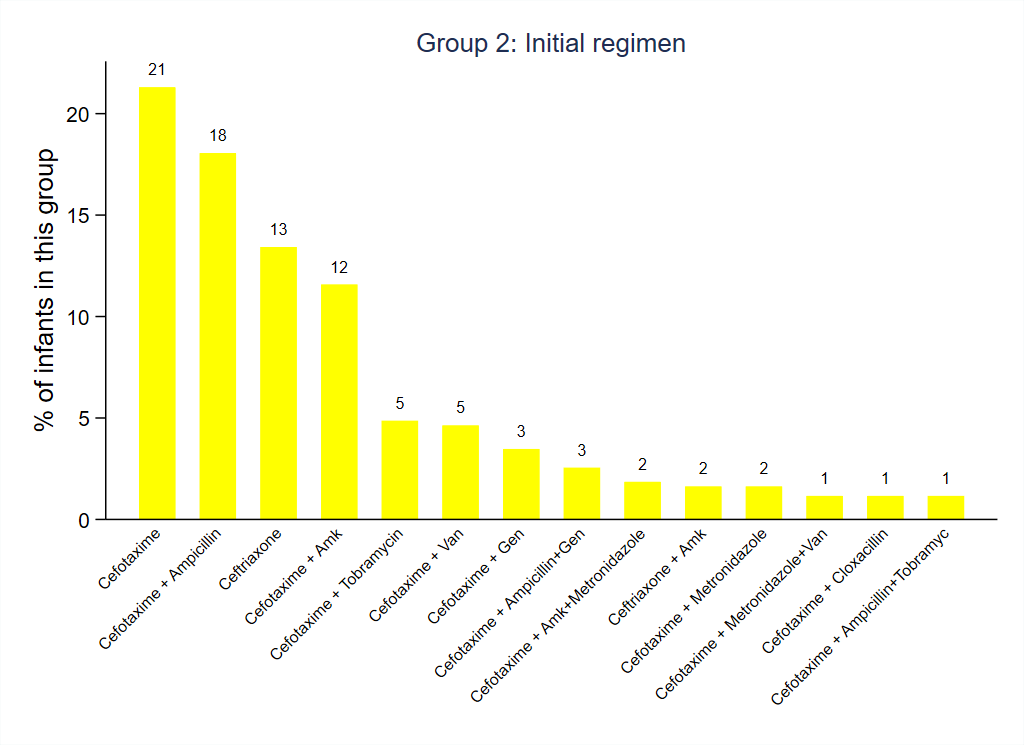

11. **Supplement figure 11: Group 3 Antibiotics - Common combinations**

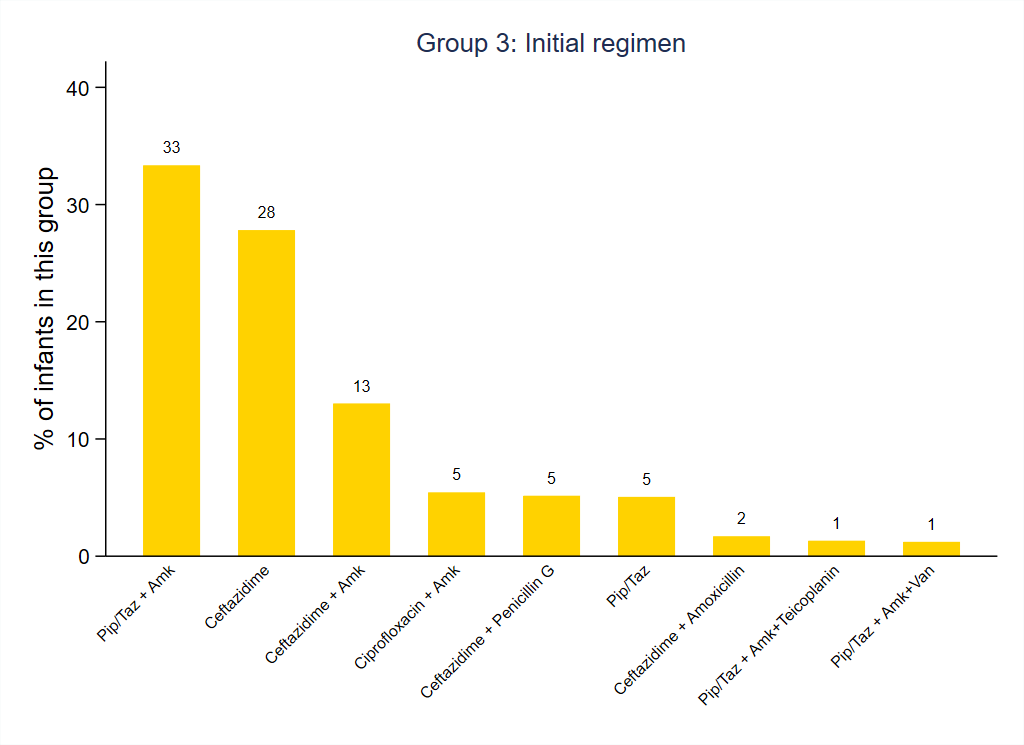

12. **Supplement figure 12: Group 4 Antibiotics - Common combinations**

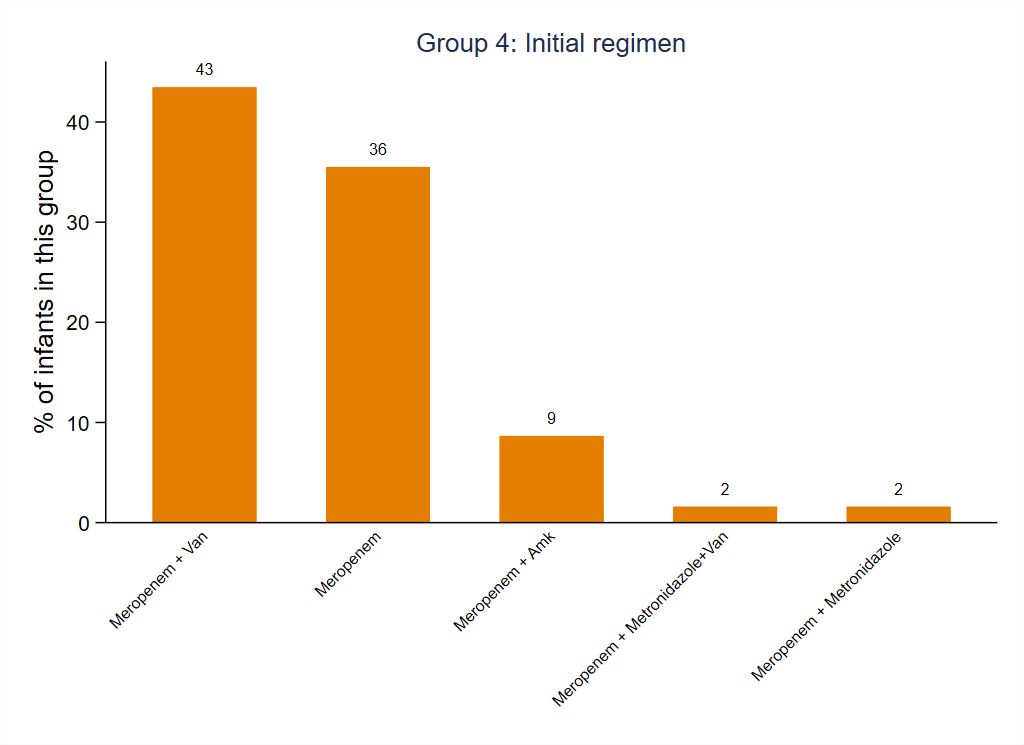

13. **Supplement figure 13: Group 5 Antibiotics - Common combinations**

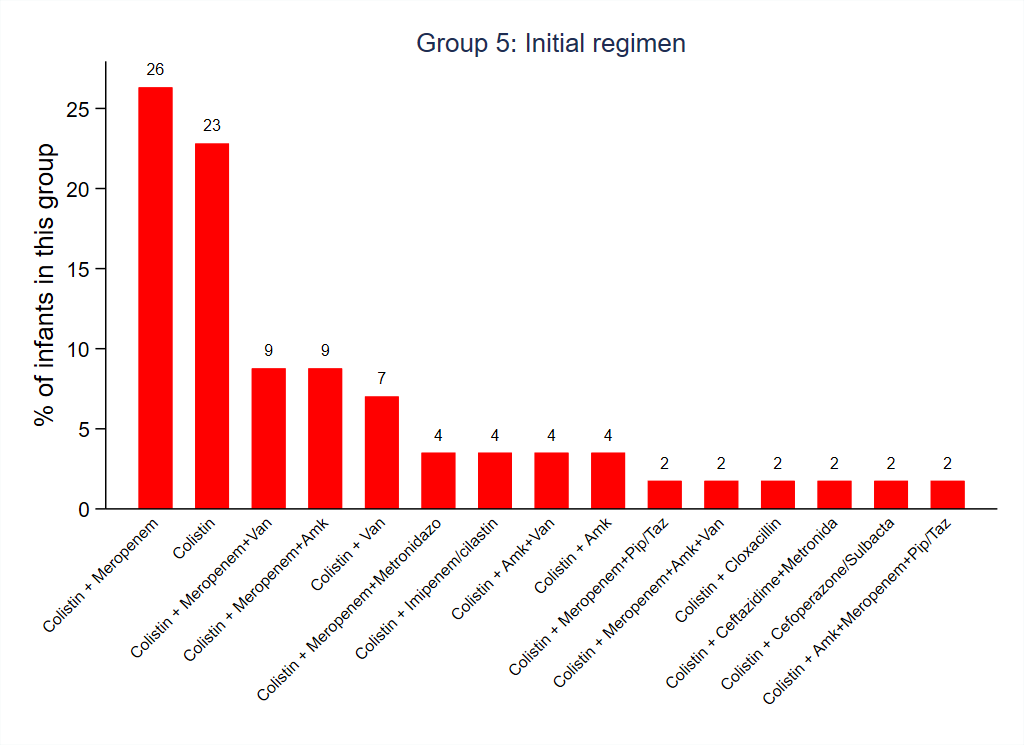

14. **Supplement figure 14: ‘Other’ Group Antibiotics - Common combinations**

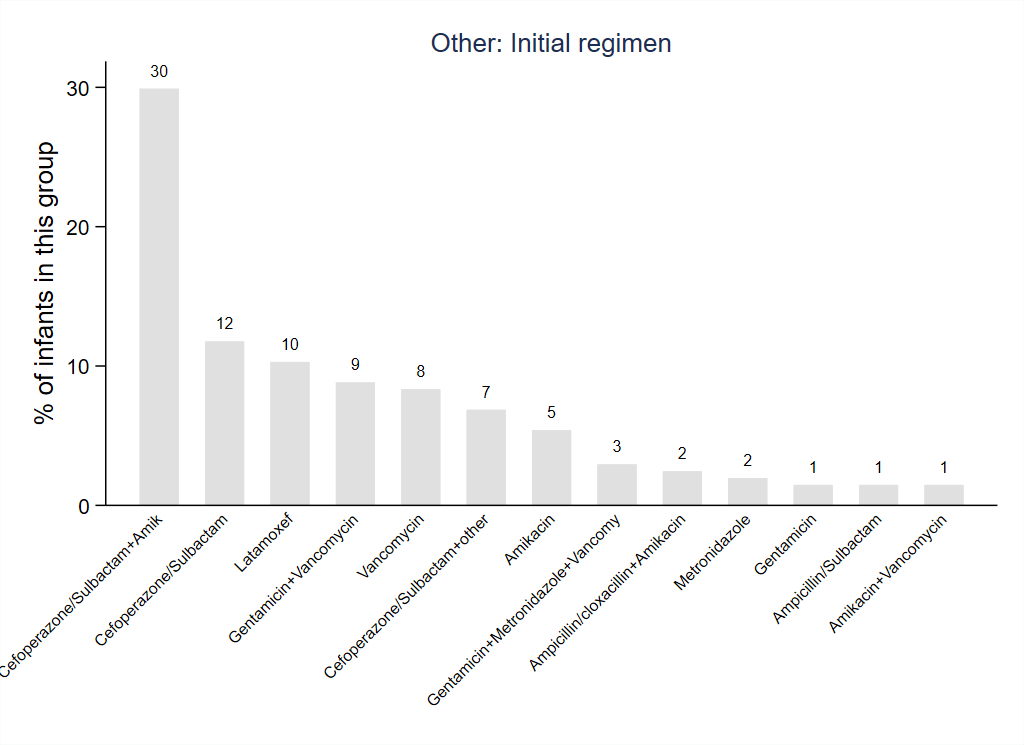

15. **Supplement figure 15: Antibiotic use by group on day 1 and day 4 post baseline**

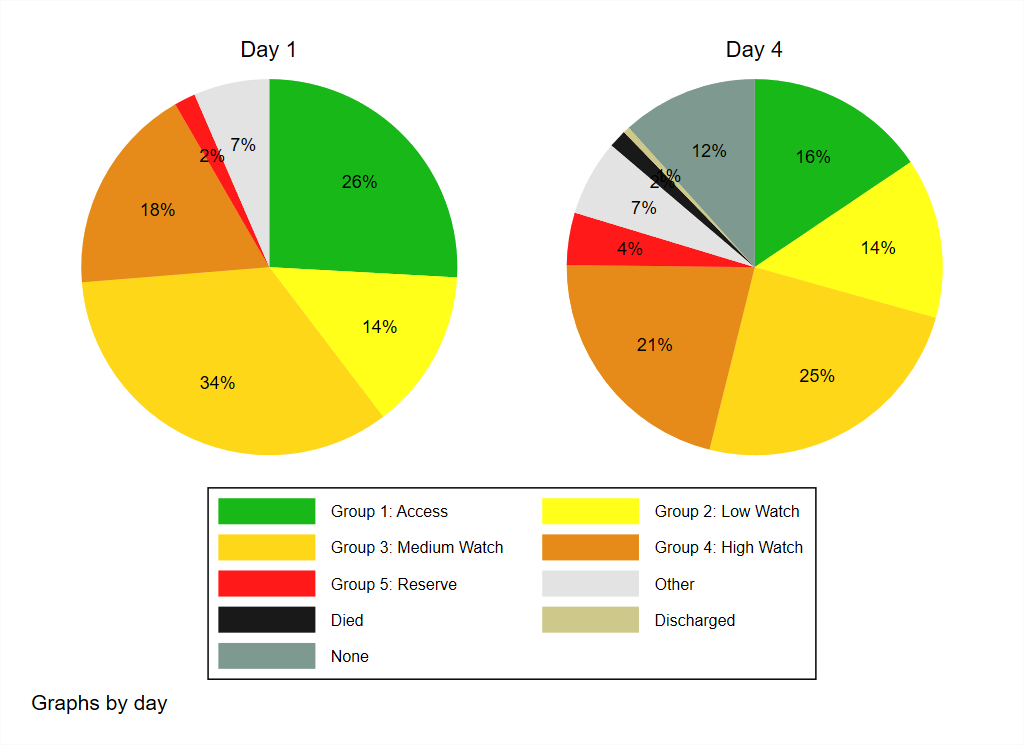

16. **Supplement figure 16: Antibiotic use by group on day 4, by initial regimen**

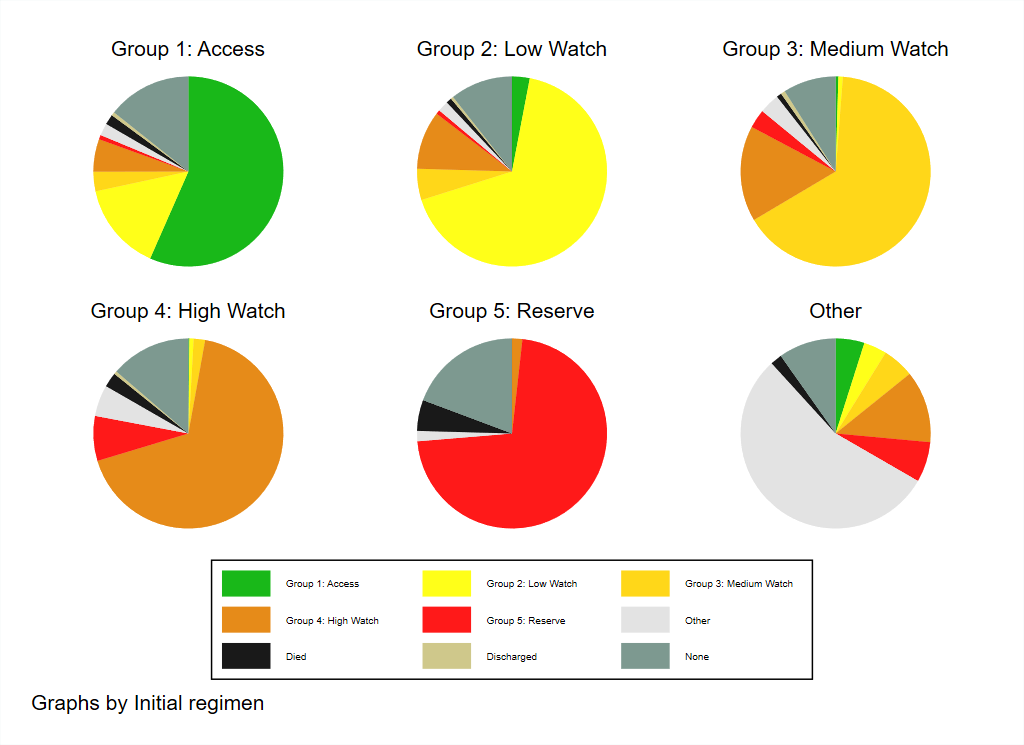

17. **Supplement figure 17: Daily antibiotic regimens, by initial regimen and overall**

| 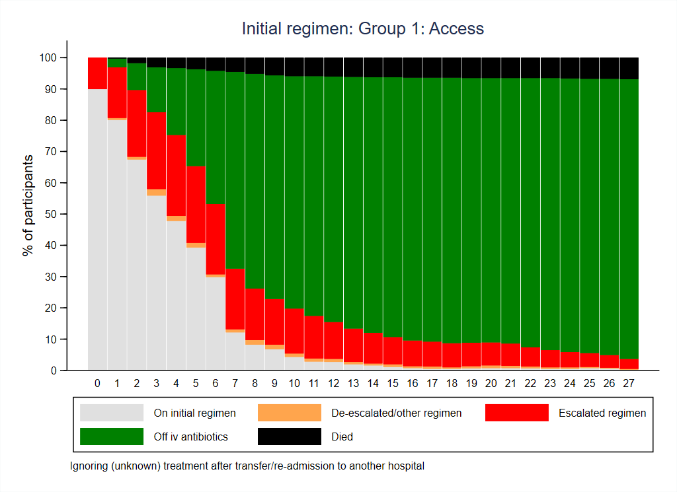 | 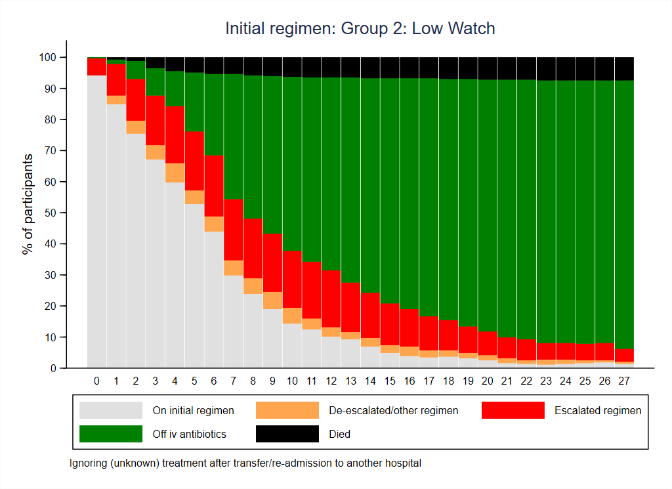 | 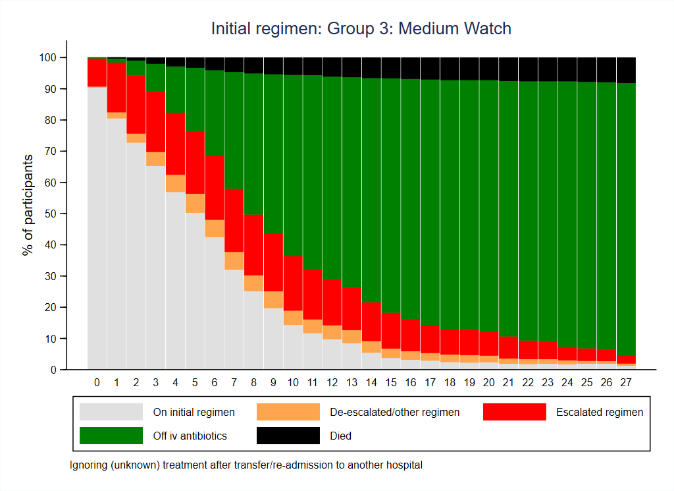 |
| --- | --- | --- |
| 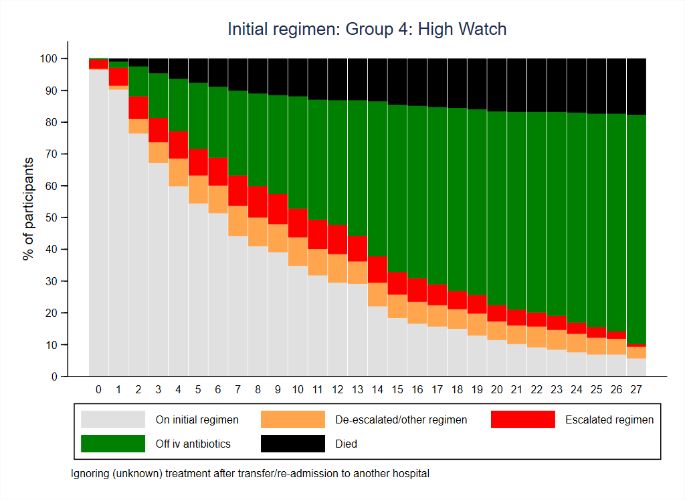 | 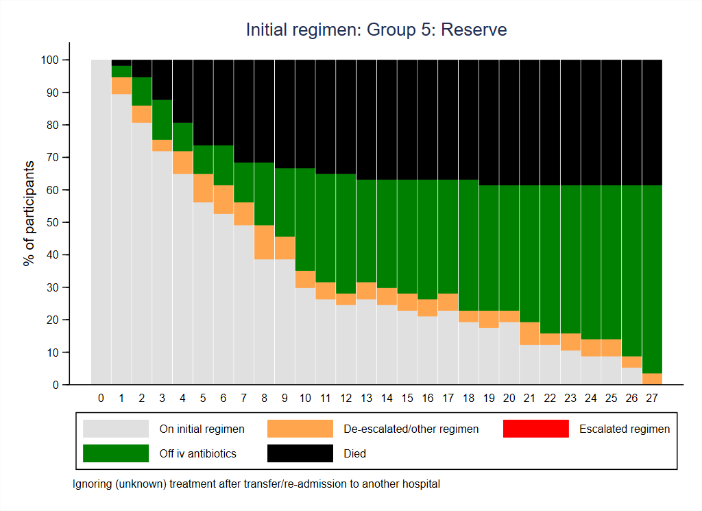 | 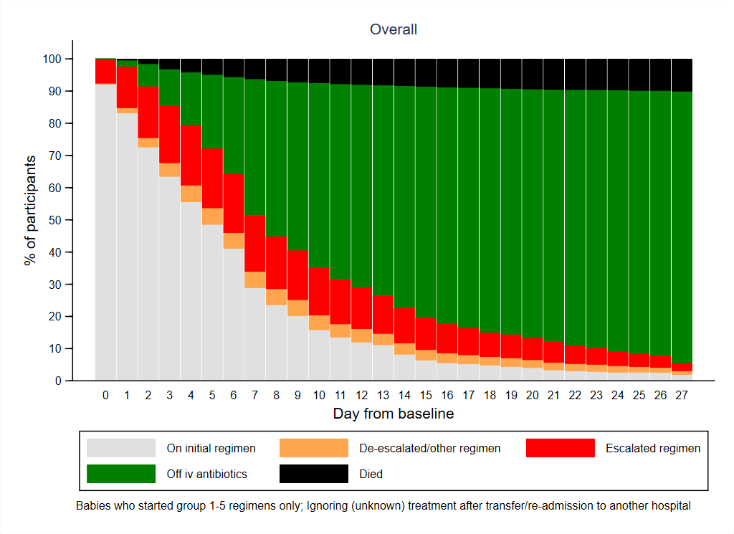 |

Note: Cross-sectional analysis.

18. **Supplement figure 18: Days on antibiotics during follow-up**

| 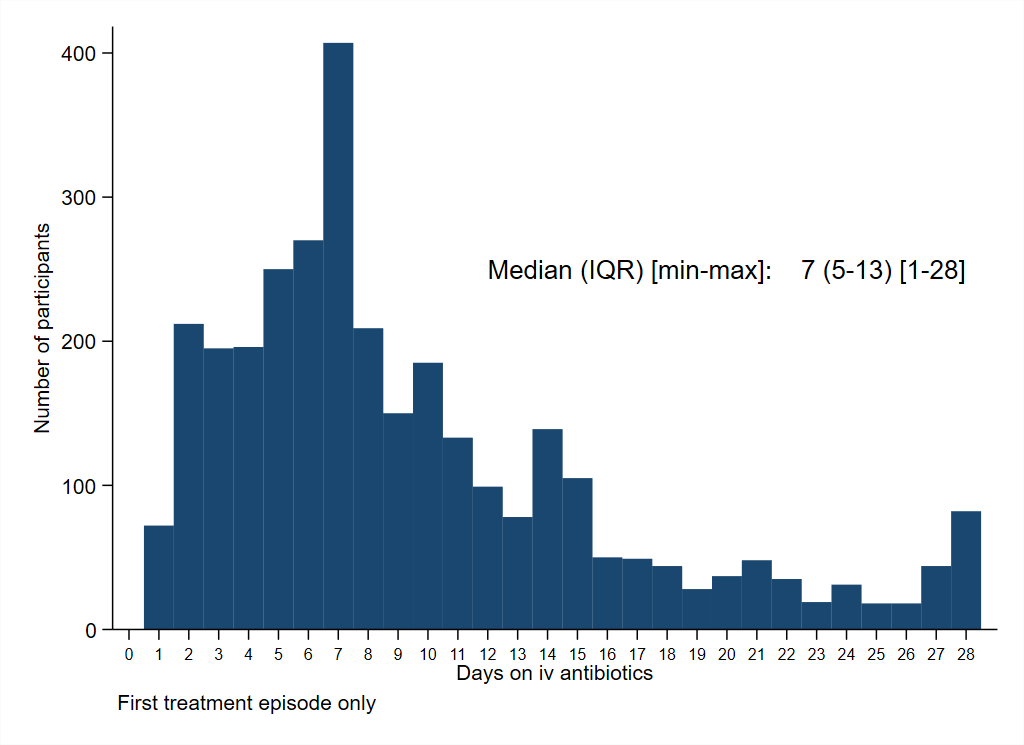 | 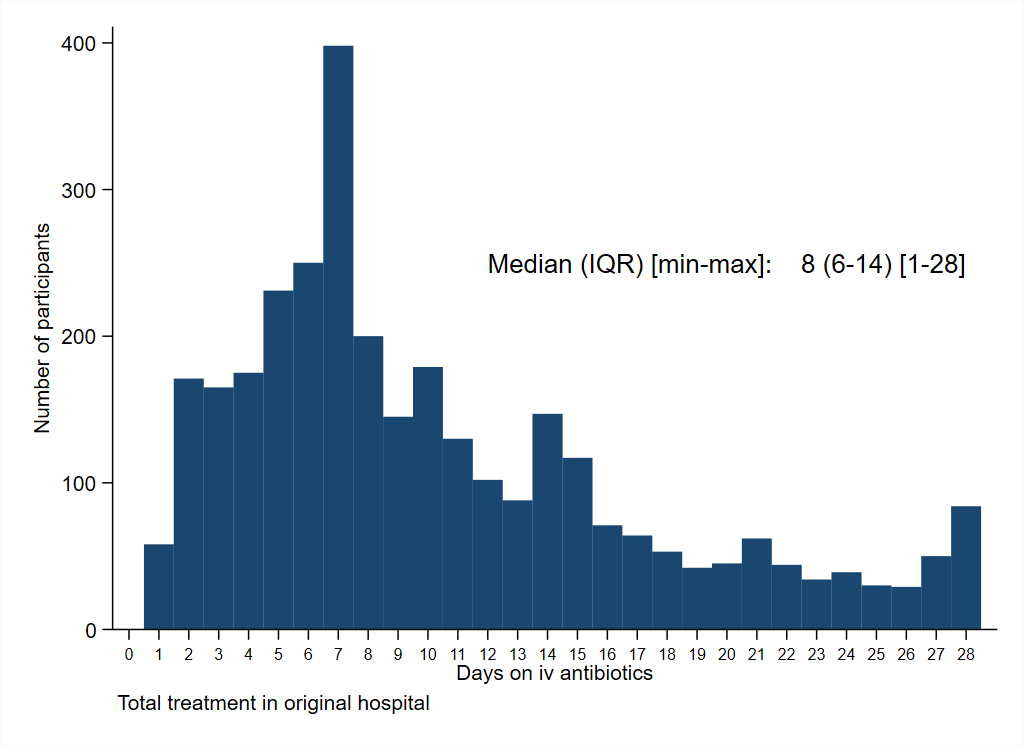 |
| --- | --- |

Note: Peak at days 27/28 due to infants still on iv antibiotics at the end of follow-up.

19. **Supplement figure 19: Frequency of organisms isolated from baseline blood culture**

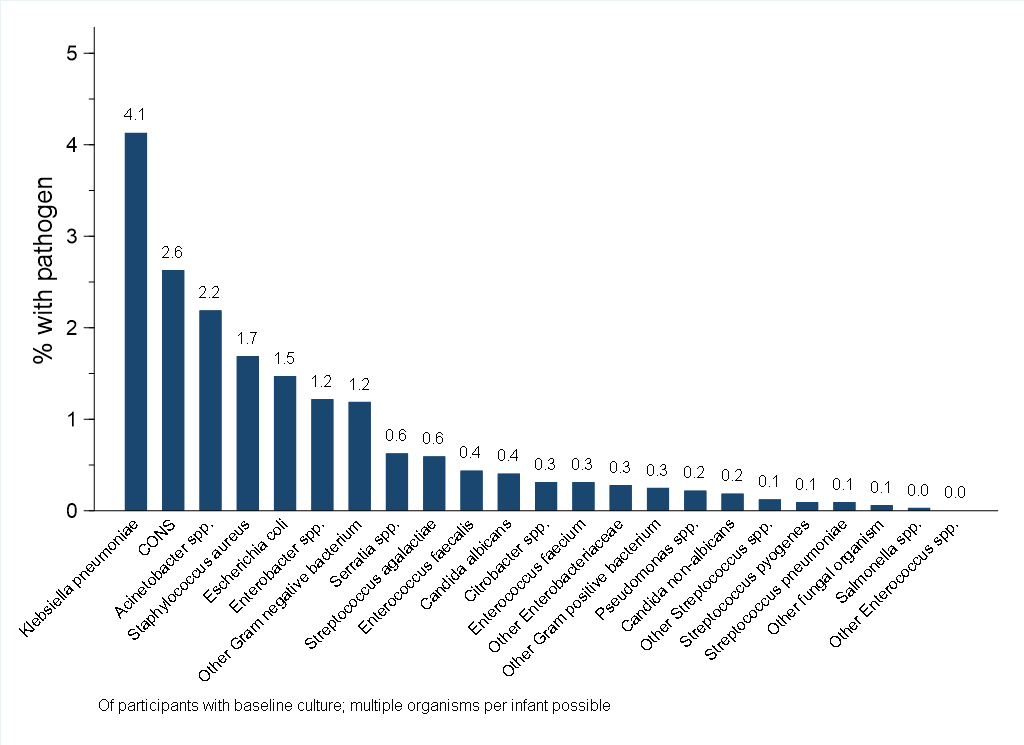

20. **Supplement figure 20: Pathogens isolated from baseline blood culture, by site**

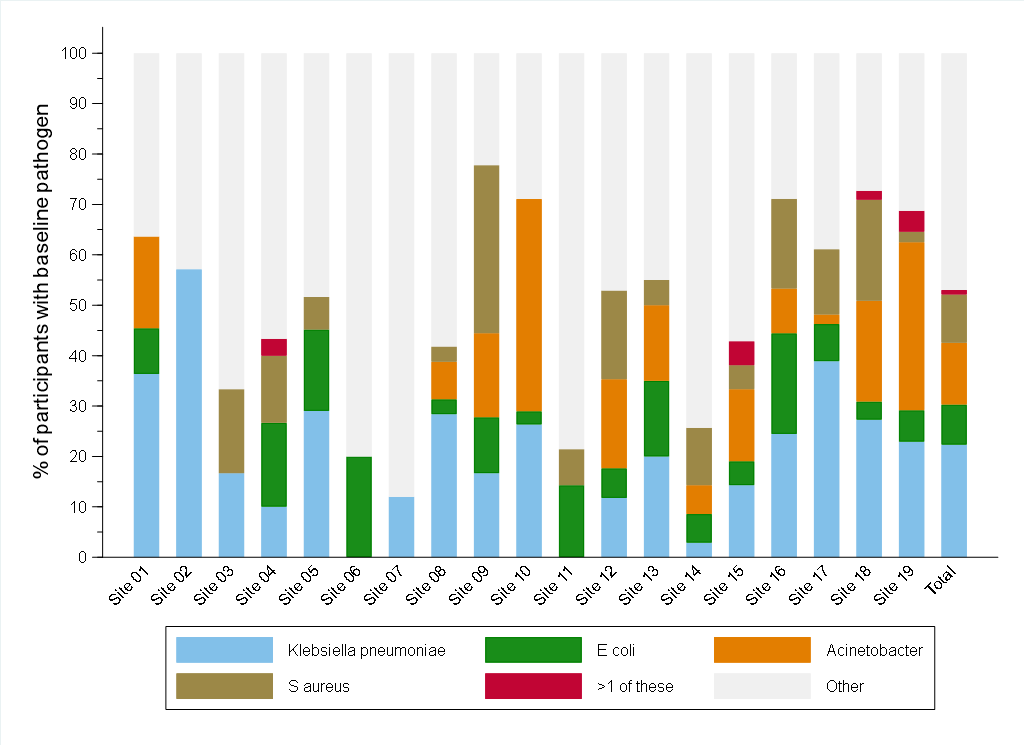

21. **Supplement figure 21: Time to death, by time to start new IV antibiotics after baseline culture**

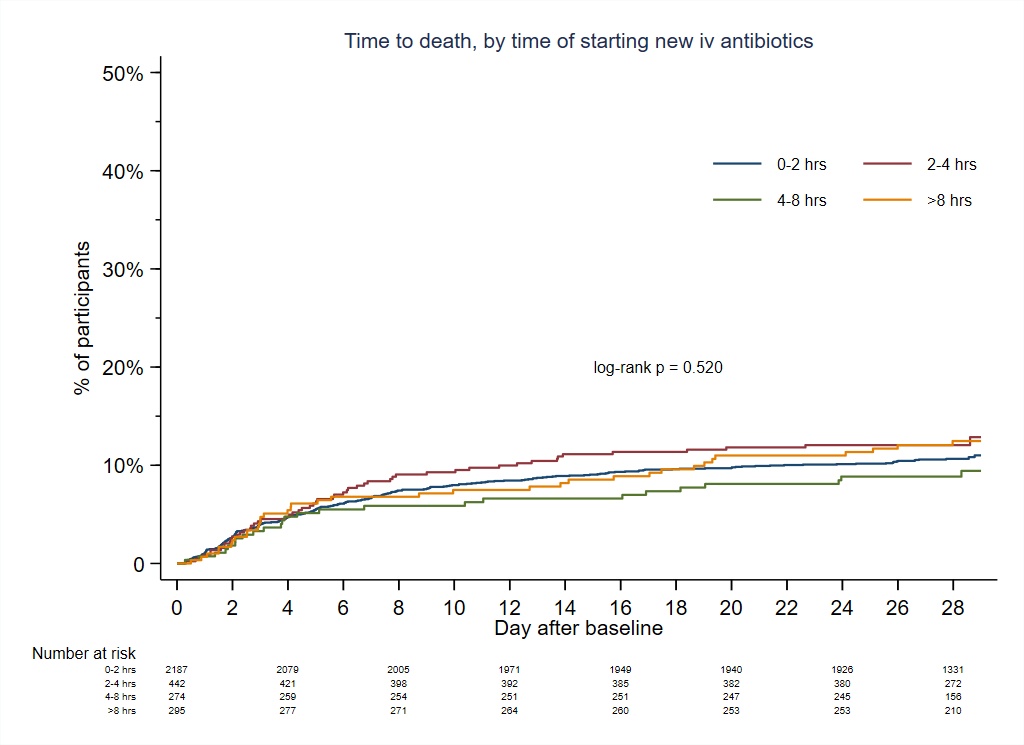

22. **Supplement table 1: Congenital anomalies**

| **Congenital anomalies** | **N=3204** |
| --- | --- |
| Does the baby have any congenital anomalies? | 265 (8.3%) |
| >1 congenital anomalies | 44 (16.6%) |
| Respiratory | 14 (0.4%) |
| Genito-urinary | 37 (1.2%) |
| Heart | 84 (2.6%) |
| Gastrointestinal system | 80 (2.5%) |
| Brain and spinal cord | 25 (0.8%) |
| Musculoskeletal | 27 (0.8%) |
| Cleft lip +/- Palate | 12 (0.4%) |
| Congenital diaphragmatic hernia | 8 (0.2%) |
| Cranio-facial | 9 (0.3%) |
| Genetic syndrome | 12 (0.4%) |
| Other | 15 (0.5%) |

23. **Supplement table 2: Laboratory results available at baseline**

|  | **N=3204** |
| --- | --- |
| White blood cell count analysed | 2800 (87.4%) |
| White blood cells (10^9/L), median (IQR) | 12.4 (7.9, 19.4) |
| Abnormal white blood cells (<4 or >20 10^9 cells/L) on day 1 |  |
| normal | 1925 (68.8%) |
| abnormal | 875 (31.3%) |
| Neutrophils analysed | 2368 (73.9%) |
| Neutrophils (10^9/L), median (IQR) | 6.8 (3.3, 12.4) |
| Neutropenia (<1.5 10^9 cells/L) on day 1 |  |
| normal | 2133 (90.1%) |
| abnormal | 235 (9.9%) |
| Platelets analysed | 2776 (86.6%) |
| Platelets (10^9/L), median (IQR) | 241.0 (158.4, 335.0) |
| Thrombocytopaenia (<150 10^9/L) |  |
| normal | 2157 (77.7%) |
| abnormal | 619 (22.3%) |
| Haemoglobin analysed | 2802 (87.5%) |
| Haemoglobin (g/dL), median (IQR) | 14.6 (12.3, 16.7) |
| Immature-to-total polymorph ratio analysed |  |
| No | 3107 (97.0%) |
| Yes | 97 (3.0%) |
| Abnormal Immature-to-total polymorph ratio on day 1 |  |
| normal | 76 (78%) |
| abnormal | 21 (22%) |
| CRP analysed | 2286 (71.3%) |
| CRP (mg/L), median (IQR) | 15 (4, 43) |
| Abnormal CRP (>10mg/L) on day 1 |  |
| Normal | 980 (42.9%) |
| abnormal | 1306 (57.1%) |
| Base excess analysed | 1492 (46.6%) |
| Base excess on day 1 |  |
| Normal | 1140 (76.4%) |
| abnormal | 352 (23.6%) |
| Lactate analysed | 1283 (40.0%) |
| Lactate on day 1 |  |
| normal | 249 (19.4%) |
| abnormal | 1034 (80.6%) |
| Acidosis analysed | 1611 (50.3%) |
| Acidosis on day 1 |  |
| normal | 467 (29.0%) |
| abnormal | 1144 (71.0%) |
| Glucose analysed | 1816 (56.7%) |
| Blood glucose (mmol/L), median (IQR) | 4.9 (3.7, 6.2) |
| Bilirubin analysed | 1187 (37.0%) |
| Blood bilirubin (µmol/L), median (IQR) | 123 (63, 184) |

24. **Supplement table 3: Initial antibiotic regimens, by time from admission**

| a) **Initial regimens for sepsis within 48hrs of admission (non-HAI)** | N=1861 | b) **Initial regimen for sepsis >48hrs after admission (HAI)** | N=1280 |
| --- | --- | --- | --- |
| Ampicillin + Gentamicin | 366 (19.7) | Meropenem + Vancomycin | 224 (17.5) |
| Ceftazidime | 248 (13.3) | Piperacillin/Tazobactam + Amikacin | 197 (15.4) |
| Piperacillin/Tazobactam + Amikacin | 159 (8.5) | Meropenem | 137 (10.7) |
| Ceftazidime + Amikacin | 102 (5.5) | Colistin (± other drug) | 55 (4.3) |
| Ampicillin + Amikacin | 86 (4.6) | Cefoperazone/Sulbactam+Amikacin | 50 (3.9) |
| Cefotaxime | 73 (3.9) | Ceftazidime | 49 (3.8) |
| Cefotaxime + Ampicillin | 69 (3.7) | Ceftazidime + Amikacin | 37 (2.9) |
| Meropenem | 64 (3.4) | Ampicillin + Gentamicin | 37 (2.9) |
| Benzylpenicillin (Penicillin G) + Gentamicin | 57 (3.1) | Meropenem + Amikacin | 34 (2.7) |
| Amoxicillin/Clavulanic acid | 44 (2.4) | Piperacillin/Tazobactam | 27 (2.1) |
| Ceftriaxone | 44 (2.4) | Amoxicillin/Clavulanic acid + Amikacin | 20 (1.6) |
| Ceftazidime + Benzylpenicillin (Penicillin G) | 44 (2.4) | Cefotaxime | 19 (1.5) |
| Ciprofloxacin + Amikacin | 40 (2.2) | Ciprofloxacin + Amikacin | 18 (1.4) |
| Amoxicillin/Clavulanic acid + Amikacin | 39 (2.1) | Ampicillin + Netilmicin | 18 (1.4) |
| Cefotaxime + Amikacin | 37 (2.0) | Gentamicin+Vancomycin | 17 (1.3) |
| Ampicillin + Tobramycin | 30 (1.6) | Vancomycin | 16 (1.3) |

25. **Supplement table 4: Factors associated with Non-WHO recommended Regimens (Groups 3-5)**

| **Factors at presentation** |  | **Odds Ratio (95% CI)** | **p-value** |
| --- | --- | --- | --- |
| Birth weight | Per additional kg | 0.57 (0.47-0.69) | <0.001 |
| Time in hospital | 0 hours  24 hours  48 hours | ref  2.28 (1.97-2.65)  4.41 (3.41-5.69) | <0.001 |
| Central line / catheter |  | 3.48 (1.74-6.94) | <0.001 |
| On iv antibiotics at baseline |  | 5.71 (3.73-8.77) | <0.001 |
| Previously positive culture |  | 25.71 (3.00-220.7) | 0.003 |
| Previous surgery |  | 5.18 (1.65-16.28) | 0.005 |
| Sepsis severity score | Per additional score point | 1.27 (1.16-1.40) | <0.001 |

Note: adjusted for centre. Non-linear relationship of time in hospital.

26. **Supplement table 5: Organisms isolated from blood at baseline, by time from admission**

| **Organisms isolated within 48hrs of admission (non-HAI)** | N=1883 | **Organisms isolated >48hrs after admission (HAI)** | N=1312 |
| --- | --- | --- | --- |
| *Escherichia coli*  *Acinetobacter* spp.  *Klebsiella pneumoniae*  *Staphylococcus aureus*  *Coagulase-negative staphylococci*  *Streptococcus agalactiae*  *Enterobacter* spp.  *Burkholderia* spp.  *Enterococcus faecalis*  *Pseudomonas* spp.  *Streptococcus pyogenes*  *Citrobacter* spp.  *Enterococcus faecium*  *Elizabethkingia anophelis*  Other* | 32 (1.7%)  21 (1.1%)  18 (1.0%)  17 (0.9%)  14 (0.7%)  13 (0.7%)  7 (0.4%)  6 (0.3%)  3 (0.2%)  3 (0.2%)  3 (0.2%)  2 (0.1%)  2 (0.1%)  2 (0.1%)  8 (0.4%) | *Klebsiella pneumoniae*  *Coagulase-negative Staphylococci*  *Acinetobacter* spp.  *Staphylococcus aureus*  *Enterobacter* spp.  *Serratia* spp.  *Elizabethkingia meningoseptica*  *Escherichia coli*  *Candida albicans*  *Enterococcus faecalis*  *Citrobacter* spp.  *Enterococcus faecium*  *Klebsiella oxytoca*  *Candida* non-*albicans*  *Streptococcus agalactiae*  *Burkholderia* spp.  *Candida non-albicans*  *Elizabethkingia anophelis*  *Bacillus* spp.  *Pseudomonas* spp.  *Streptococcus pneumoniae*  Other* | 114 (8.7%)  70 (5.3%)  51 (3.9%)  37 (2.8%)  32 (2.4%)  19 (1.5%)  15 (1.1%)  15 (1.1%)  13 (1.0%)  11 (0.8%)  8 (0.6%)  8 (0.6%)  7 (0.5%)  6 (0.5%)  6 (0.5%)  6 (0.5%)  6 (0.5%)  5 (0.4%)  4 (0.3%)  4 (0.3%)  2 (0.2%)  12 (0.9%) |

Note: * Organisms isolated in only one infant per group.

27. **Supplement table 6: Susceptibility result for pathogens in baseline blood culture.**

|  |  | **Klebsiella pneumoniae** | **Acinetobacter spp.** | **Escherichia coli** | **Staphylococcus aureus** | **Serratia spp.** | **Burkholderia spp.** | **Elizabethkingia meningoseptica** |
| --- | --- | --- | --- | --- | --- | --- | --- | --- |
| **ampicillin** | S | 0 | 0 | 0 | 21 (39% | 0 | 0 | 0 |
|  | R | 132 (100%) | 72 (100%) | 47 (100%) | 33 (61%) | 20 (100%) | 12 (100%) | 15 (100%) |
| **gentamicin** | S | 50 (38%) | 19 (27%) | 37 (80%) | 24 (57%) | 18 (90%) | 0 | 0 |
|  | I | 5 (4%) | 2 (3%) | 0 | 3 (7%) | 0 | 0 | 0 |
|  | R | 75 (58%) | 50 (70%) | 9 (20%) | 15 (36%) | 2 (10%) | 12 (100%) | 15 (100%) |
| **ceftriaxone** | S | 29 (23%) | 0 | 27 (63%) | 21 (39%) | 7 (64%) | 0 | 0 |
|  | I | 2 (2%) | 0 | 0 | 0 | 0 | 0 | 0 |
|  | R | 96 (76%) | 72 (100%) | 16 (37%) | 33 (61%) | 4 (36%) | 12 (100%) | 15 (100%) |
| **cefotaxime** | S | 30 (23%) | 0 | 29 (64%) | 21 (39%) | 7 (64%) | 0 | 0 |
|  | I | 2 (2%) | 0 | 0 | 0 | 0 | 0 | 0 |
|  | R | 96 (75%) | 72 (100%) | 16 (36%) | 33 (61%) | 4 (36%) | 12 (100%) | 15 (100%) |
| **ceftazidime** | S | 26 (28%) | 0 | 29 (78%) | 0 | 7 (64%) | 12 (100%) | 0 |
|  | I | 2 (2%) | 0 | 0 | 0 | 0 | 0 | 0 |
|  | R | 66 (70%) | 72 (100%) | 8 (22%) | 54 (100%) | 4 (36%) | 0 | 15 (100%) |
| **ciprofloxacin** | S | 51 (50%) | 14 (24%) | 11 (44%) | 21 (53%) | 13 (93%) | 1 (13%) | 4 (31%) |
|  | I | 4 (4%) | 0 | 2 (8%) | 0 | 1 (7%) | 2 (25%) | 9 (69%) |
|  | R | 48 (47%) | 45 (76%) | 12 (48%) | 19 (48%) | 0 | 5 (63%) | 0 |
| **levofloxacin** | S | 20 (56%) | 13 (48%) | 10 (53%) | 16 (64%) | 4 (80%) | 3 (27%) | 13 (100%) |
|  | I | 2 (6%) | 3 (11%) | 3 (16%) | 0 | 1 (20%) | 1 (9%) | 0 |
|  | R | 14 (39%) | 11 (41%) | 6 (32%) | 9 (36%) | 0 | 7 (64%) | 0 |
| **piperacillin-tazobactam** | S | 50 (44%) | 0 | 38 (90%) | 21 (39%) | 6 (75%) | 0 | 1 (7%) |
|  | I | 11 (10%) | 0 | 2 (5%) | 0 | 1 (13%) | 0 | 0 |
|  | R | 53 (46%) | 72 (100%) | 2 (5%) | 33 (61%) | 1 (13%) | 12 (100%) | 13 (93%) |
| **meropenem** | S | 88 (67%) | 19 (27%) | 38 (97%) | 21 (39%) | 18 (90%) | 10 (91%) | 0 |
|  | I | 1 (1%) | 1 (1%) | 0 | 0 | 0 | 1 (9%) | 0 |
|  | R | 43 (33%) | 50 (71%) | 1 (3%) | 33 (61%) | 2 (10%) | 0 | 15 (100%) |
| **colistin** | S | 39 (100%) | 43 (98%) | 4 (100%) | 0 | 0 | 0 | 0 |
|  | I | 0 | 1 (2%) | 0 | 0 | 0 | 0 | 0 |
|  | R | 0 | 0 | 0 | 54 (100%) | 19 (100%) | 12 (100%) | 15 (100%) |
| **amikacin** | S | 80 (62%) | 19 (27%) | 43 (93%) | 24 (57%) | 15 (75%) | 0 | 0 |
|  | I | 10 (8%) | 1 (1%) | 1 (2%) | 3 (7%) | 1 (5%) | 0 | 0 |
|  | R | 40 (31%) | 51 (72%) | 2 (4%) | 15 (36%) | 4 (20%) | 12 (100%) | 15 (100%) |
| **vancomycin** | S | 0 | 0 | 0 | 42 (100%) | 0 | 0 | 0 |
|  | R | 132 (100%) | 72 (100%) | 47 (100%) | 0 | 20 (100%) | 12 (100%) | 15 (100%) |
| **methicillin** | S | 31 (23%) | 0 | 24 (51%) | 21 (39%) | 1 (5%) | 0 | 0 |
|  | I | 0 | 0 | 1 (2%) | 0 | 0 | 0 | 0 |
|  | R | 101 (77%) | 0 | 22 (47%) | 33 (61%) | 19 (95%) | 0 | 0 |

Note: S=susceptible; I=intermediate; R=resistant. Denominators across drugs variables due to available susceptibility results.

28. **Supplement table 7: Organisms isolated from CSF in first 7 days from baseline**

| **Class** | **Organism** | **Pathogen** | **Contaminant** | **Unclassified** |
| --- | --- | --- | --- | --- |
| Gram-positive | *Streptococcus agalactiae* | 3 | 0 | 0 |
|  | *Coagulase-negative Staphylococci* | 2 | 7 | 1 |
|  | *Enterococcus faecalis* | 2 | 1 | 0 |
|  | *Enterococcus faecium* | 2 | 0 | 0 |
|  | *Bacillus* spp. | 0 | 12 | 1 |
|  | *Micrococcus* spp. | 0 | 1 | 0 |
|  | *Streptococcus oralis* | 0 | 1 | 0 |
|  | *Corynebacterium* spp. | 0 | 1 | 0 |
| Gram-negative | *Acinetobacter baumannii* | 11 | 0 | 0 |
|  | *Escherichia coli* | 9 | 0 | 0 |
|  | *Klebsiella pneumoniae* | 9 | 0 | 0 |
|  | *Elizabethkingia meningoseptica* | 4 | 0 | 0 |
|  | *Elizabethkingia anophelis* | 3 | 0 | 0 |
|  | *Enterobacter* spp. | 2 | 0 | 0 |
|  | *Burkholderia* spp. | 1 | 0 | 0 |
|  | *Proteus mirabilis* | 1 | 0 | 0 |
|  | *Sphingomonas paucimobilis* | 1 | 0 | 0 |
|  | *Acinetobacter ursingii* | 0 | 1 | 0 |

Note: n=73 with positive CSF culture in first 7 days.

29. **Supplement table 8: Pathogens and mortality**

| **Factor** | **Univariable** | **Adjusted for unmodifiable baseline predictors^*^** | **Adjusted for all baseline predictors^**^** |
| --- | --- | --- | --- |
| Pathogen at baseline | 1.65 (1.30 – 2.10)  *p<0.001* | 1.54 (1.21 – 1.97) *p=0.001* | 1.57 (1.22-2.02)  *p<0.001* |
| Pathogen type at baseline No pathogen Gram positive pathogen Gram negative pathogen Fungal pathogen | ref. 0.96 (0.57 – 1.62) 1.80 (1.37 – 2.35) 4.55 (2.21 – 9.36)  *p<0.0001* | ref. 1.06 (0.63 – 1.80) 1.62 (1.24 – 2.12) 3.84 (1.79 – 8.20)  *p=0.0003* | ref.  1.35 (0.79-2.30)  1.55 (1.17-2.04)  4.24 (1.94-9.30)  *p=0.0007* |
| Pathogen at baseline No pathogen Acinetobacter Klebsiella pneumoniae E. coli S. aureus Streptococcus agalactiae Other pathogen**^***^** | ref. 2.70 (1.76 – 4.14) 1.92 (1.29 – 2.86) 1.77 (0.79 – 4.00) 1.03 (0.46 – 2.33) 2.65 (0.98 – 7.18) 1.17 (0.80 – 1.73)  *p=0.0001* | ref. 2.01 (1.30 – 3.10) 1.77 (1.18 – 2.66) 2.13 (0.94 – 4.81) 1.02 (0.45 – 2.31) 3.30 (1.20 – 9.03) 1.14 (0.77 – 1.69)  *p=0.0032* | ref.  1.96 (1.27-3.05)  1.49 (0.99-2.25)  2.33 (1.01-5.37)  1.22 (0.53-2.80)  3.40 (1.22-9.48)  1.28 (0.86-1.92)  *p=0.0098* |

Note: Results are HR (95% CI). All models are adjusted for site (random effect). **^*^** adjusted for birth weight, gestational age, time in hospital, congenital anomalies, and site; **^**^** adjusted for all factors in NeoSep Severity Score (as in ^*^ plus abdominal distension, difficulty in feeding, evidence of shock, lethargy/no movement, temperature, level of respiratory support); **^***^** including CONS.
